# Pathway-based integration of multi-omics data reveals lipidomics alterations validated in an Alzheimer’s Disease mouse model and risk loci carriers

**DOI:** 10.1101/2021.05.10.21255052

**Authors:** Monica Emili Garcia-Segura, Brenan R. Durainayagam, Sonia Liggi, Gonçalo Graça, Beatriz Jimenez, Abbas Dehghan, Ioanna Tzoulaki, Ibrahim Karaman, Paul Elliott, Julian L. Griffin

## Abstract

Alzheimer’s Disease (AD) is a highly prevalent neurodegenerative disorder. Despite increasing evidence of important metabolic dysregulation in AD, the underlying metabolic changes that may impact amyloid plaque formation are not understood, particularly for late onset AD. This study analyzed genome-wide association studies (GWAS), transcriptomics and proteomics data obtained from several data repositories to obtain differentially expressed (DE) multi-omics elements in mouse models of AD. We characterized the metabolic modulation in these datasets using gene ontology, and transcription factor, pathway and cell-type enrichment analysis. A predicted lipid signature was extracted from genome-scale metabolic networks (GSMN) and subsequently validated in a lipidomic dataset derived from cortical tissue of ABCA7-null mice, a mouse model of one of the genes associated with late onset AD. Moreover, a metabolome-wide association study (MWAS) was performed to further characterize the association between dysregulated lipid metabolism in human blood serum and AD.

We found 203 DE transcripts, 164 DE proteins and 58 DE GWAS-derived mouse orthologs associated with significantly enriched metabolic biological processes. Lipid and bioenergetics metabolic pathways were significantly over-represented across the AD multi-omics datasets. Microglia and astrocytes were significantly enriched in the lipid-predominant AD-metabolic transcriptome. We also extracted a predicted lipid signature that was validated and robustly modelled class separation in the ABCA7 mice cortical lipidome, with 11 of these lipid species exhibiting statistically significant modulations. MWAS revealed 298 AD single nucleotide polymorphisms (SNP)-metabolite associations, of which 70% corresponded to lipid classes.

These results support the importance of lipid metabolism dysregulation in AD and highlight the suitability of mapping AD multi-omics data into GSMNs to identify metabolic alterations.

## 1 Introduction

Alzheimer’s Disease (AD) is a neurodegenerative disorder prevalent in later life characterized by amyloid deposition, hyperphosphorylated tau aggregation into neurofibrillary tangles and a sustained neuroinflammatory response (DeTure & Dickson 2019). With the proportion of the population over 65 years of age increasing annually, a mechanistic understanding of the disease is urgently needed (Xie *et al*. 2020). There are several emerging lines of evidence highlighting the importance of metabolic dysfunctions in AD. Impaired glycolysis and bioenergetics shifts towards fatty-acid and amino-acid metabolism seem to indicate that mitochondrial dysfunction or substrate switch play a role in AD pathogenesis (Perez Ortiz & Swerdlow 2019). Cholesterol metabolism can also exert lipotoxic effects in the AD brain via ceramide production modulation (Cutler *et al*. 2004). Furthermore, there are several genes linked to AD onset and progression that are also related to brain lipid metabolism. The apolipoprotein epsilon4 (APOE4) allele, the strongest risk factor for AD development, is known to cause significant disruptions in brain lipid homeostasis in both human carriers and transgenic animals (Fernandez *et al*. 2019). Similarly, triggering receptor expressed on myeloid cells-2 (TREM2), another gene strongly associated with AD, actively undergoes lipid-sensing and consequently induces changes in the microglia lipidome (Nugent *et al*. 2020). Finally, loss-of-function variant of the ATP - binding-casette, subfamily-A, member-7 gene (ABCA7) has been strongly associated with late-onset AD (De Roeck *et al*. 2019). ABCA7 has been implicated in AD pathology through amyloid-precursor protein (APP) endocytosis, impaired amyloid-beta (Aβ) clearance and, although not fully elucidated, lipid metabolism dysregulation via sterol regulatory element binding protein 2 (SREBP2) (Aikawa *et al*. 2018).

Despite all the accumulating evidence, mechanistic explanations of AD have mostly been centered around amyloid or tau-centric hypotheses, and therefore much remains to be understood regarding the underlying metabolic processes (Johnson *et al*. 2020). Multi-omics approaches have the potential to overcome the limitations of the current knowledge in this field. These approaches can provide a comprehensive view of a particular pathophysiological state by interrogating molecular changes across several levels of biological functions (Canzler *et al*. 2020). A promising methodological approach relevant to the study of metabolites is genome scale metabolic networks (GSMN), which uses genomics and transcriptomics data to predict metabolic pathway modulations (Pinu *et al*. 2019). GSMN also allow for the interpretation of multi-omics data via metabolic subnetwork curation, thus providing an attractive metabolic framework which can be effectively validated using metabolomics and lipidomics data (Frainay & Jourdan 2017).

The aim of this study was to validate the presence of metabolic perturbations in AD using multi-omics pathway-based integration and extraction of metabolic subnetworks from open source data (**Figure 1**). We found consistent perturbations of lipid and energy metabolism across three AD multi-omics datasets compiled from previous studies, from which we extracted 133 lipid species predicted to be dysregulated in AD which we then validated in an ABCA7 knock-out (KO) mouse dataset acquired with ultraperformance liquid chromatography-mass spectrometry (UPLC-MS). The importance of this association was explored further by performing a metabolome-wide association study (MWAS) of the blood plasma metabolome for AD risk loci carriers in two human cohorts using ^1^H NMR spectroscopy.

**Figure 1.**
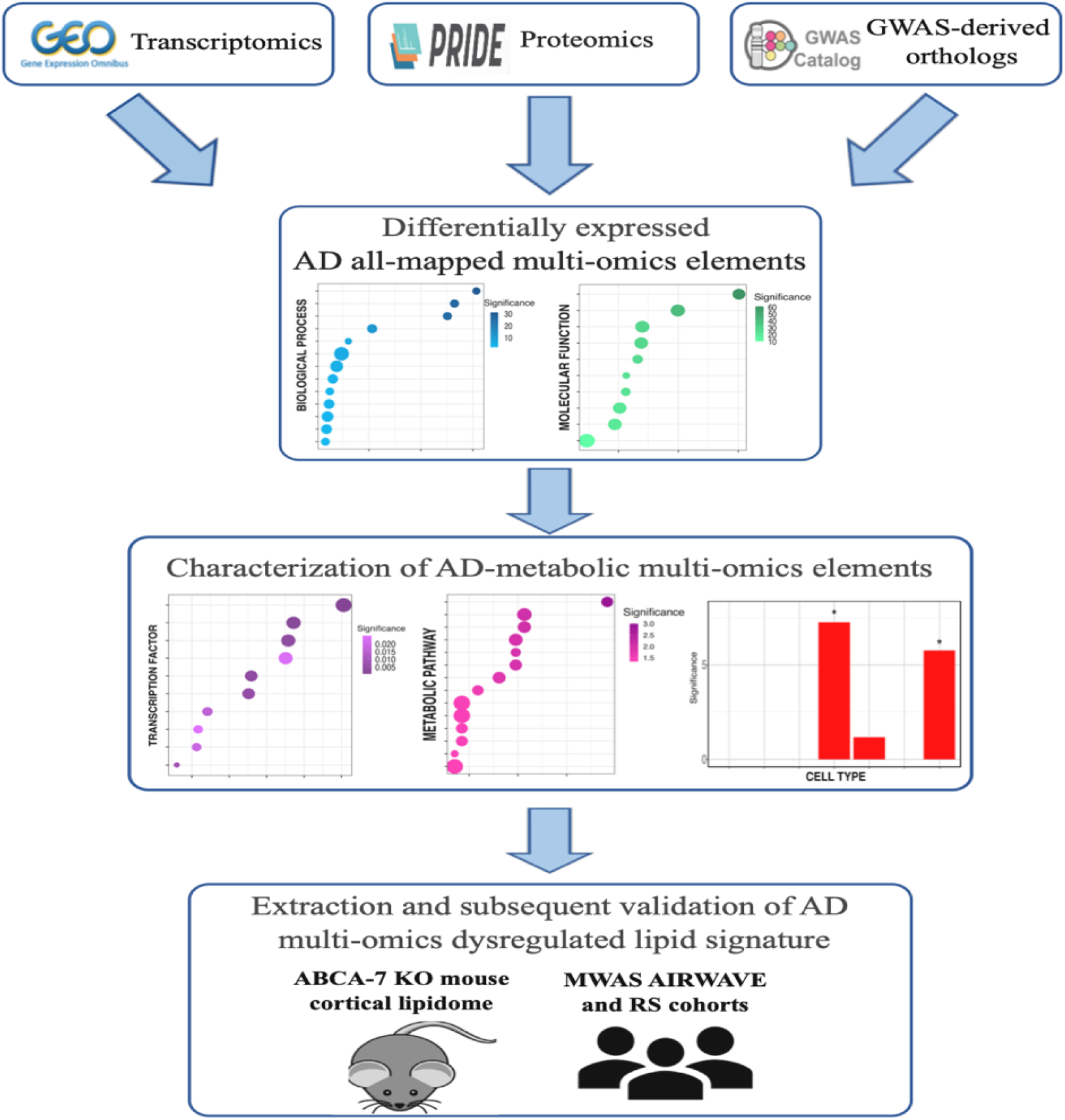
Schematic representation of the experimental design implemented in this study. Abbreviations: GEO = gene expression omnibus database, PRIDE = protein identification database, AD = Alzheimer’s Disease, ABCA7 KO = ATP-binding-cassette, subfamily A, member 7 gene knock-out.

This study also highlights the suitability of interpreting multi-omics data in the context of GSMNs, as the predicted lipid terms and species were not only found in the cortical ABCA7 lipidome, but its associated multivariate model robustly separated ABCA7 mice from their wild-type (WT) litter-mates.

## 2 Materials and Methods

### 2.1 Data collection of AD mouse brain transcriptomics and proteomics data

The gene expression omnibus (GEO) repository (https://www.ncbi.nlm.nih.gov/geo/) (Clough & Barrett 2016) was queried on 15/06/20 for gene expression studies using “Alzheimer’s Disease” as our search term. The following criteria were employed for dataset selection: *Mus musculus* organism, expression profiling by array as study-type, tissue as attribute, brain tissue expression compared to WTs and a minimum of 3 animals per condition. This search yielded 11 datasets (GSE25926, GSE53480, GSE60460, GSE77574, GSE77373, GSE109055, GSE111737, GSE113141, GSE141509 and GSE74441) from 9 studies (Aydin *et al*. 2011; Polito *et al*. 2014; Hamilton *et al*. 2015; Marsh *et al*. 2016; Wang *et al*. 2017; Faivre *et al*. 2018; Hou *et al*. 2018; Fang *et al*. 2019; Preuss *et al*. 2020). The proteomics identifications (PRIDE) repository (Jones *et al*. 2006) was queried on 01/07/20 for proteomics studies applying the following filters: Alzheimer’s Disease as disease, brain as organism-part and *Mus musculus* as organism. Datasets comparing the AD proteome against WTs, with minimum 3 animals per condition and with deposited proteinGroups.txt files were included. This search yielded 4 datasets (PXD007795, PXD011068, PXD012238, and PXD007813) from 4 publications (Palomino-Alonso *et al*. 2017; Hamezah *et al*. 2019; Kim *et al*. 2019; Lachen-Montes *et al*. 2019). However, differences in protein expression failed to reach statistical significance after controlling for the false discovery rate (FDR) in two studies (Palomino-Alonso *et al*. 2017; Hamezah *et al*. 2019), and thus their corresponding datasets were excluded. A description of all included datasets can be found in **Table 1**.

**Table 1.** Characteristics of the transcriptomics and proteomics datasets included in this study

| Brain region | GEO/PRIDE<br>accession number | AD<br>animal model | Age | Sample size | Platform |
| --- | --- | --- | --- | --- | --- |
| **Transcriptomics** |  |  |  |  |  |
| **Frontal cortex** | GSE113141 | APP/PS1 | 9-10 months | AD (n=6)<br>WT (n=6) | Agilent-074809 SurePrint<br>G3 Mouse GE v2 8x60K<br>Microarray |
|  | GSE109055 | 3xTgAD | 22-24<br>months | AD (n=4)<br>WT(n=4) | Agilent-028005 SurePrint<br>G3 Mouse GE 8x60K<br>Microarray |
|  | GSE77373 | 5xFAD | 5 months | AD (n=3)<br>WT(n=3) | Affymetrix Mouse Gene<br>1.0 ST Array |
|  | GSE74441 | APP/PS1 | Not<br>disclosed | AD (n=6)<br>WT (n=6) | Illumina MouseRef-8 v2.0<br>expression beadchip |
|  | GSE25926 | APP-KI | 24-28<br>months | AD (n=3)<br>WT(n=3) | Affymetrix Mouse<br>Genome 430 2.0 Array |
| **Hippocampus** | GSE111737 | APP/PS1 | 8 months | AD (n=6)<br>WT (n=6) | Agilent-074809 SurePrint<br>G3 Mouse GE v2 8x60K<br>Microarray |
|  | GSE109055 | 3xTgAD | 22-24<br>months | AD (n=4)<br>WT(n=4) | Agilent-028005 SurePrint<br>G3 Mouse GE 8x60K<br>Microarray |
|  | GSE53480 | Tg4510 | 4 months | AD (n=4)<br>WT(n=4) | Affymetrix Mouse<br>Genome 430 2.0 Array |
| **Subventricular zone** | GSE60460 | 3xTgAD | 7 months | AD (n=4)<br>WT(n=4) | Agilent-028005 SurePrint<br>G3 Mouse GE 8x60K<br>Microarray |
| **Half-brain** | GSE141509 | 5xFAD | 6 months | AD (n=6)<br>WT (n=6) | NanoString nCounter®<br>Mouse AD panel |
| **Whole brain** | GSE77574 | 5xFAD | 6-7 months | AD (n=4)<br>WT(n=4) | Affymetrix Mouse<br>Transcriptome Array 1.0 |
| **Proteomics** |  |  |  |  |  |
| **Hippocampus** | PXD012238 | 5xFAD | 10 months | AD (n=6)<br>WT (n=6) | Orbitrap MS/MS- Q-<br>Exactive |
| **Olfactory bulb** | PXD007813 | Tg2576 | 6 months | AD (n=3)<br>WT(n=3) | iTRAQ-LC MS/MS<br>with Triple TOF MS 5600 |

### 2.2 Differential expression (DE) analysis of AD mouse transcriptomics and proteomics data

Processed transcriptomics datasets were retrieved from GEO using the GEOquery Bioconductor-based package (version 2.54.1) (Davis & Meltzer 2007) in the R environment, version 3.6.2 (https://www.R-project.org). Datasets were log-2 transformed and graphically inspected to verify appropriate data normalization; probes that were not mapped to any genes, mapped to more than one gene and probes with missing values (N/As) were filtered out. Differential expression analysis was performed using significance analysis of microarray (SAM) with *samr* package (version 3.0) (Tusher *et al*. 2001) within the R environment. SAM can control for the total number of false positives through both gene specific t-tests and a maximum local tolerable FDR (Tusher *et al*. 2001). Upon 200 permutation-based SAM analysis, multiple testing correction was applied by adjusting the total false positives to 3% and the local FDR for 90^th^ percentile of DE genes to 5% in every dataset.

Proteomics datasets were analyzed using Perseus (version 1.6.5) (Tyanova *et al*. 2016). Initially, proteins only identified by reverse-decoy, site or known contaminants were excluded, as well as proteins with 2/3 of replicates per group reporting N/As. Protein intensities were then log-2 transformed and remaining N/As were replaced using normal distribution values, as most proteomics studies assume N/As are indicative of low-expression proteins (Tyanova *et al*. 2016). DE proteins were determined using a two-tailed Student’s t-test with a 200 FDR permutation-based method and a 0.050 p-value cut-off (Tusher *et al*. 2001). In isobaric tag for relative and absolute quantification (iTRAQ) experiments, an additional fold change (FC) 1.17-0.83 cut-off was introduced to determine DE proteins. iTRAQ experiments are prone to interference/ratio distortion (Pappireddi *et al*. 2019), and thus a combination of p-value, FDR and FC cut-off is the most suitable approach to detect biological variability (Oberg & Mahoney 2012).

### 2.3 AD genome-wide association studies (GWAS) gene-based analysis and mouse ortholog determination

AD GWAS summary statistics were obtained from a meta-analysis of the UK-Biobank and International Genomics of Alzheimer’s Project (IGAP) cohorts, which evaluated GWAS with AD by-proxy in 388364 individuals across both cohorts (Marioni *et al*. 2018). Summary statistics (ID: GCST005922) were retrieved from the NHGRI-EBI GWAS-Catalog (https://www.ebi.ac.uk/gwas/) (Buniello *et al*. 2019) on 07/07/2020.

Gene-based analysis was performed with multi-marker analysis of genomic annotation (MAGMA, version 1.07bb) (de Leeuw *et al*. 2015), using gene locations from the genome reference consortium-human build-37 (GRCh37, NCBI) and a reference panel of European ancestry from the 1000 genomes project phase-3 (Auton *et al*. 2015). MAGMA provides a combined p-statistic of genes significantly associated with single nucleotide polymorphisms (SNPs) (de Leeuw *et al*. 2015); we used a combined 0.050 p-value as a significance cut-off. Significant genes were imported into Ensembl –Biomart on 20/07/2020 (version GrCh37.13; https://grch37.ensembl.org/biomart/martview) to determine high confidence mouse orthologs (Zerbino *et al*. 2018). Upon excluding genes associated with either several or no mouse orthologs, only those exhibiting one-to-one bidirectional orthology with 60% protein sequence similarity across both species were considered high-quality mouse orthologs (Mancuso *et al*. 2019).

### 2.4 Gene ontology (GO) analysis and AD-metabolic multi-omics extraction

DE transcripts, proteins and GWAS-orthologs were initially mapped onto the BioCyc *Mus musculus* GSMN (Caspi *et al*. 2016) using MetExplore, which provides a framework for metabolic subnetwork extraction(Cottret *et al*. 2018). DE transcripts, protein-coding and GWAS-orthologs genes that were not mapped onto the GSMN were removed; the resulting omics lists are referred to as “all-mapped” data throughout this study. Significantly enriched functional terms were identified in all-mapped AD omics datasets using the database for annotation, visualization and integrated discovery (DAVID, version 6.8) (https://david,neifcrf.gov/) (Dennis *et al*. 2003). and the *Mus musculus* genome as background. GO analysis was performed using a hypergeometric test with an EASE score of 0.1 and a count threshold of 2. Terms with both raw p-value and Benjamini-Hochberg (B-H) FDR-adjusted p-value (α) below 0.050 were considered statistically significant. Metabolism - related transcripts, proteins and GWAS-orthologs were manually extracted from significantly enriched biological processes (BP).

### 2.5 Transcription Factor (TF) enrichment analysis

TF enrichment analysis was performed on all-mapped AD genes and proteins, as well as their metabolic counterparts, using ChIP-X enrichment analysis 3 (ChEA3) (https://maayanlab.cloud/chea3/). ChEA3 performs enrichment analysis based on TF’s target genes coverage using the Fishers exact test and B-H adjusted p-value at 0.050 threshold (Keenan *et al*. 2019). The ENCODE library was chosen as our reference set, as it incorporates TF-target associations from human and mouse data (Davis *et al*. 2018). Significantly enriched TF were manually cross-referenced with the mouse transcription factor atlas to verify its mouse tissue expression (Zhou *et al*. 2017).

### 2.6 Pathway enrichment analysis of AD-metabolic multi-omics data

AD metabolic transcripts, proteins and GWAS-orthologs lists were mapped onto the BioCyc *Mus musculus* GSMN (Caspi *et al*. 2016) in MetExplore (Cottret *et al*. 2018). Metabolic pathway enrichment analysis was performed using hyper-geometric tests with right-tailed Fishers exact tests with B-H correction for multiple testing (α=0.050).

### 2.7 Expression-weighted cell-type enrichment (EWCE) of AD-metabolic multi-omics data

EWCE was conducted on AD-metabolic transcriptomics, proteomics and GWAS-orthologs datasets using the *EWCE* package in R (version 0.99.2)(Skene & Grant 2016). EWCE computes an enrichment p-value that describes the probability of an input gene list having a meaningful expression within a specific cell-type upon 10000 random permutations (Skene & Grant 2016). A cortical and hippocampal single-cell RNA-sequencing dataset with large coverage was used as background (Zeisel *et al*. 2015); B-H adjusted p-values were calculated using the R base package. A conditional EWCE analysis was also performed on the combined AD-metabolic multi-omics dataset to probe the relationships between enriched cell-types, using an approach originally developed for GWAS data analysis (Skene *et al*. 2018).

### 2.8 Metabolic subnetwork extraction

To ultimately validate lipid alterations highlighted during pathway enrichment analysis, a metabolic subnetwork containing all lipid terms or species in significantly enriched lipid pathways was mined across the AD-metabolic transcriptome and proteome using MetExplore (Cottret *et al*. 2018). After excluding non-lipid metabolites, a combined predicted lipid signature across the AD multi-omics datasets was created, which was visualized using MetExploreViz (Chazalviel *et al*. 2018). Lipid identifiers were then retrieved from LIPIDMAPS (Fahy *et al*. 2009).

### 2.9 Cortical ABCA7-KO lipidomics dataset

We also employed a lipidomics dataset of cortical extracts of 7 WT and 7 ABCA7-KO 11-months old mice, with 3 females and 4 males per group, as described previously (Aikawa *et al*. 2018). Lipidomic extraction was performed on ∼50mg cortex tissue using a modified Folch extraction (Su *et al*. 2019). Global lipidomic profiling of the cortical extracts and 3 pooled samples was acquired using a reverse-phase ultraperformance liquid chromatography - mass spectrometry (RP-UPLC-MS) on a Synapt Quadruple-Time of Flight mass spectrometer (Waters Corp., Manchester, UK) in positive and negative mode. Details of systems configuration and analytical conditions have been previously reported (Andreas *et al*. 2020). Data processing was performed with KniMet (Liggi *et al*. 2018). Briefly, signals extracted using the R library XCMS (Tautenhahn *et al*. 2012) were retained if present in at least 50% of the pooled samples with a Coefficient of Variation <= 20. Remaining signals were subjected to imputation of N/As using K-Nearest Neighbour (KNN), probabilistic quotient normalization (PQN) based on pooled samples, and annotation using LIPID MAPS (https://lipidmaps.org/; (Fahy *et al*. 2009)), retention time matching to standards and fragmentation data.

### 2.10 Multivariate statistical analysis

Multivariate statistical analysis was performed on both positive and negative mode for the original ABCA7-KO and validated lipid signature subsets using P-SIMCA (Umetrics, Sweden) following log-transformation of intensities and Pareto-scaling. Orthogonal projections to latent structures-discriminant analysis (OPLS-DA) models, which allow to evaluate the impact of group membership by separating the variance attributed or orthogonal to class membership into components, were created for both original datasets and validated subset in positive and negative ion mode (Griffin *et al*. 2020). Lipids in the validated subset in positive and negative mode with variable influence of projection (VIP) > 1 were retained for univariate analysis, as OPLS-DA generated VIP > 1 indicate specific variables with important contributions to the model (Liu *et al*. 2020). The suitability of the models were assessed through inspection of their R^2^(cum)X and Q^2^ values, which respectively represent the percentage of model-captured variation and predictive capability (Liu *et al*. 2020). Models were further validated with a 100 permutation-based test, in which the correlation coefficient for the permuted class-membership variable is plotted against the R ^2^(cum)X and Q^2^(cum) (Murgia *et al*. 2017).

### 2.11 Univariate statistical analysis

AD multi-omics lipid species that had an associated VIP score above 1 in the original ABCA7 KO lipidomics dataset underwent univariate statistical analysis using GraphPad Prism (p<0.05). Negative-mode acquired lipids underwent both a Student t-test and Mann-Whitney non-parametric test comparing genotype (p<0.05), whereas positive-mode acquired lipids were analyzed using One-way ANOVA comparing genotype and sex correcting for multiple testing using B-H method (α<0.05).

### 2.12 Metabolome-wide association study (MWAS) of the blood plasma metabolome for AD risk loci carriers

We performed an MWAS using nuclear magnetic resonance (NMR) spectra of blood from 3258 individuals from the Airwave Health Monitoring Study (Airwave) and the Rotterdam Study (RS) prospective cohorts (Elliott *et al*. 2014; Ikram *et al*. 2020). Ethical approval for access to the Airwave cohort was granted following application to the access committee via the Dementia Platform UK (https://portal.dementiasplatform.uk/). Access to the RS cohort was granted following access to the Management Committee and conducted under approval from the Ministry of Health, Welfare and Sport of the Netherlands. Blood samples were heparin plasma for Airwave and serum for RS. Average age at enrolment in 2004 was 40.9 years for men and 38.5 years for women in the Airwave cohort; the RS cohort mean age of recruitment was 55 for both genders in 1990 (Elliott *et al*. 2014; Ikram *et al*. 2020)

Sample preparation and metabolic profiling in these cohorts have been extensively described (Tzoulaki *et al*. 2019; Robinson *et al*. 2020). Briefly, ^1^H NMR solvent suppression pulse and T2-Carr-Purcell-Meiboom-Gill (CPMG) spectra were acquired per sample (Dona A.C. *et al*. 2014) and additionally lipid quantification was applied on the ^1^H NMR solvent suppression pulse spectra using a commercial package (Jiménez *et al*. 2018). Resonances associated with both protons attached to the fatty acid and the head group (largely choline and glycerol) along with protons from cholesterol and cholesterol esters were classified as belonging to the lipid class.

MWAS was performed using 47 unique genetic loci based on three recent GWAS meta-analysis on AD to identify AD risk loci carriers (Lambert *et al*. 2013; Jansen *et al*. 2019; Kunkle *et al*. 2019). These studies evaluated genome-wide associations with late-onset AD (LOAD) in individuals across the IGAP and UK-Biobank cohorts.

### 2.13 MWAS association statistics

We carried out a linear regression to calculate the effect estimates of each SNP with all metabolomic features (23,571 data points for original NMR spectra and 105 features for the fitted lipid data) with adjustment for age, sex, and cohort. Prior to the analysis, each cohort data was residualised using 10 principal components from genome-wide scans to adjust for population stratification. To account for multiple testing, we used a permutation-based method to estimate the Metabolome Wide Significance Level (MWSL) to consider the high degree of correlation in metabolomics datasets (Chadeau-Hyam M *et al*. 2010; Castagné R *et al*. 2017). A P-value threshold giving a 5% Family-Wise Error Rate was computed for each SNP in each data platform.

## 3 Results

### 3.1 DE analysis of mapped AD mouse transcriptomics and proteomics data

DE transcripts and proteins in the AD mouse brain with potential metabolic functions were extracted from the GEO and PRIDE repositories, respectively. Microarray expression profiles from 11 datasets were obtained from 5 distinct brain regions (frontal cortex, hippocampus, sub-ventricular zone, brain hemisphere and whole-brain) and 5 AD mouse models (APP/PS1, 5xFAD, 3xTgAD, APP-KI and Tg4510; **Table 1**). SAM revealed 2884 DE genes with a 90^th^ percentile FDR below 5%. Of these, 594 were accurately mapped onto the GSMN, which were used to generate the all-mapped AD transcriptomics dataset. Furthermore, proteomics datasets from the hippocampus and olfactory bulb of 5xFAD and Tg2576 mice, respectively, were also obtained (**Table 1**). Permutation-based analysis revealed 1537 DE proteins (FDR p < 0.050), of which 392 were mapped onto the GSMN and therefore constituted the all - mapped AD proteomics dataset. DE proteins from two additional studies (Palomino-Alonso *et al*. 2017; Hamezah *et al*. 2019) failed to reach statistical significance upon FDR correction and thus these datasets were removed from further analysis.

### 3.2 Mapped high-quality mouse orthologs identification from gene-based AD GWAS analysis

High-confidence mouse orthologs of significantly associated genes in human AD GWAS studies were also identified to gain a more comprehensive view of metabolic perturbations in AD. Gene-based analysis with MAGMA (de Leeuw *et al*. 2015) using summary statistics from 388364 individuals in the UK-Biobank and IGAP cohorts (Marioni *et al*. 2018) revealed 18178 gene-level associations with human AD SNPs, of which 1664 were considered significant (combined p-value < 0.05). After applying high-quality mouse orthology criteria (Mancuso *et al*. 2019), 1356 high-quality orthologs of AD SNPs-associated human genes were identified. The all-mapped AD GWAS-orthologs dataset was generated by accurately mapping 258 GWAS-orthologs onto the GSMN.

### 3.3 Differential GO and TF enrichment analysis across AD multi-omics datasets

Potential TF and GO enrichment were investigated across the AD multi-omics datasets. More than 25% of mapped AD protein-coding genes were also found in the AD transcriptomics dataset (**Figure 2A**). In terms of up-stream regulation, 67 TF were significantly enriched in the all-mapped AD proteome, whereas only 17 TF were enriched in the all-mapped AD transcriptome (**Table S1**). Despite these differences, *CCCTC-binding factor* (*CTCF), TAL BHLH transcription factor 1* (*TAL1), MYC associated factor X* (*MAX)* and *basic helix-loop-helix family member E40* (*BHLHE40)* were among the top10 potential enriched TFs across both datasets (FDR p<0.050, **Figure 2B, Table S1)**.

**Figure 2.**
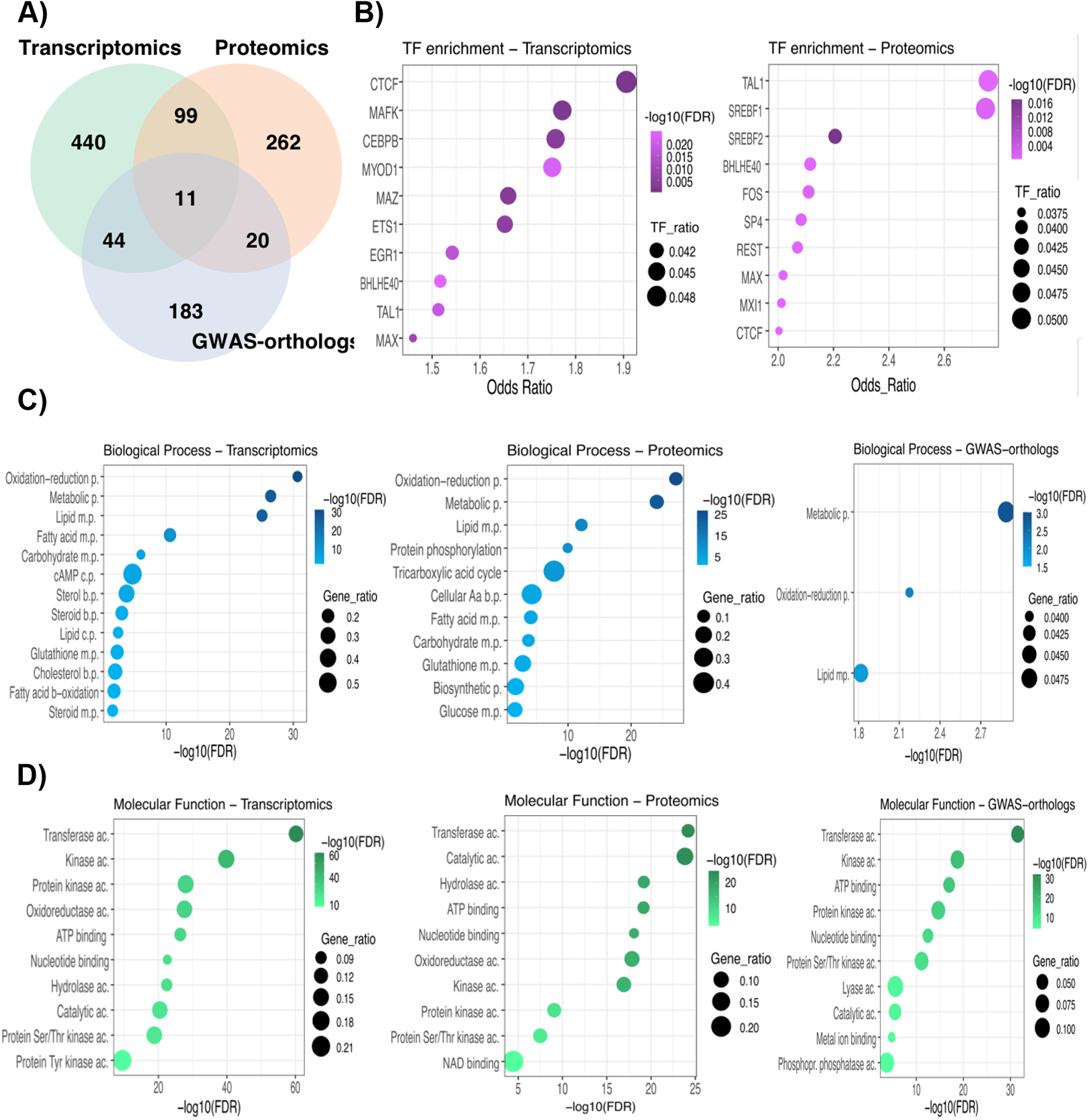
Transcription factor and functional enrichment analysis reveal shared functional processes between all-mapped AD multi-omics datasets. **(A)** Venn Diagram showing the amount of overlap between AD mapped transcripts, proteomics and GWAS - orthologs genes. (**B)** Top 10 TF enrichment analysis results of AD transcriptomics and proteomics datasets. (**C)** Selected Biological Process (BP) functional enrichment analysis of three AD multi-omics datasets. “M.p”, “b.p.” and “c.p.” refer to metabolic, biosynthetic and catabolic processes, respectively. (**D)** Top 10 Molecular (MF) functional enrichment analysis of three AD multi-omics datasets. “Ac” refers to molecular function activity. TF ratio refers to the number of mapped input genes in relation to the total TF’s target genes. –log10(FDR) refers to the inverse, log-transformed FDR-adjusted enrichment p-value. Gene ratio refers to the number of mapped input genes in relation to all Gene Ontology (GO) term-associated genes. The entire list of over-represented TF and GO terms can be found in **Table S1 and S2**, respectively.

GO analysis revealed shared functional terms across the three datasets (**Figure 2C-D**). Oxidation-reduction, lipid and fatty-acid metabolic processes were enriched in all-mapped AD transcriptomics and proteomics (FDR p<0.050, **Figure 2C**). Six additional lipid-related BP terms were over-represented in all-mapped AD transcriptomics data, whereas the TCA cycle was only enriched in the AD proteome (FDR p<0.050, **Figure 2C**). Transferase, catalytic, ATP binding, kinase activity, nucleotide binding and serine/threonine-kinase activity were among the top10 over-represented terms across all-mapped AD multi-omics datasets (FDR p<0.050, **Figure 2D**). Cytosol and mitochondria were the cellular compartment (CC) terms most over-represented in the all-mapped AD transcriptome and proteome respectively; membrane was the only significant CC term in the AD GWAS - orthologs dataset (**Table S2**).

### 3.4 Lipid-related metabolic pathways and regulators are enriched across AD-metabolic multi-omics datasets

Given the elevated number of metabolic BP significantly enriched across the three multi-omics datasets, the DE 203 transcripts, 164 proteins and 58 GWAS-orthologs genes mapped to these BP were subjected to further characterization. The largest degree of overlap was again found between AD-metabolic transcripts and proteins (**Figure 3A**). Although there were substantially more enriched TFs in the AD-metabolic proteome (**Table S3**), lipid-associated TFs such as *estrogen-related receptor alpha (ESRRα)* and *sterol regulatory element binding transcription factor 1* (*SREBF1)* were overrepresented in the AD-metabolic transcriptome and proteome (FDR p<0.050, **Figure 3B**). Pathway enrichment analysis reflected differential metabolic processes across the multi-omics datasets (**Figure 3C**). Pathways related to cholesterol, phospholipases and fatty-acid metabolism were significantly over-represented in the AD-metabolic transcriptomics dataset, whereas the AD-metabolic proteome was associated with mitochondrial processes such as TCA cycle, glycolysis and NADH electron transfer (FDR p<0.050, **Figure 3C**). Lipid processes such as CPD-diacylglycerol and phosphatidylglycerol synthesis were also enriched in AD-metabolic proteome (**Figure 3C**). Thyroid hormone metabolism was significantly enriched in the GWAS-orthologs dataset with 66% pathway coverage (**Table S4**).

**Figure 3.**
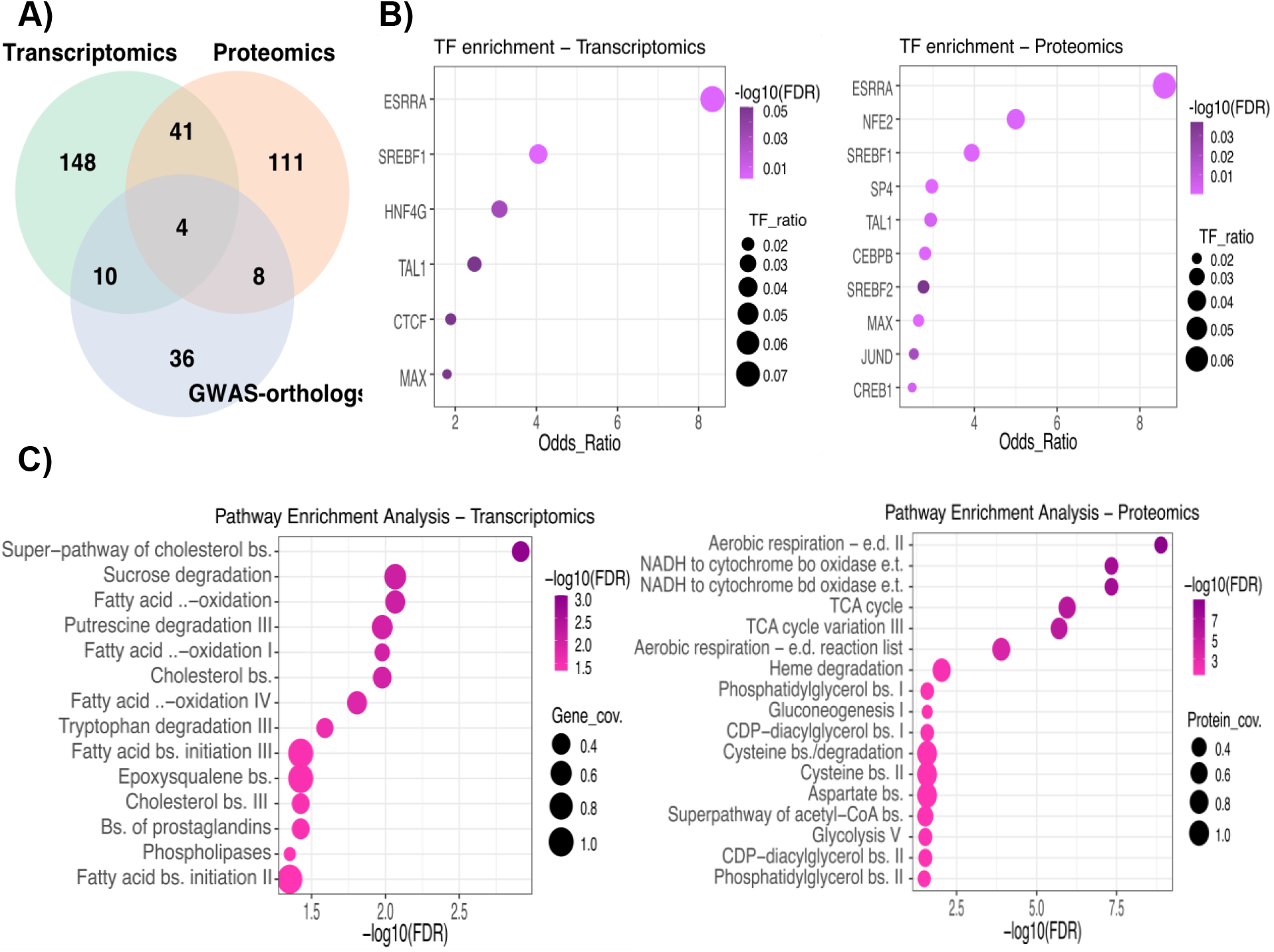
TF and pathway enrichment analysis highlights enrichment of lipid-related metabolic processes in metabolic transcriptomic and proteomic datasets from mouse models of AD. **(A)** Venn Diagram showing the amount of overlap between AD metabolic multi-omics datasets. (**B)** Top 10 TFs significantly overrepresented in AD metabolic transcripts and proteins. (**C)** Pathway enrichment analysis of the three AD multi-omics datasets. “bs.”, “e.d.” and “e.t.” refer to biosynthesis, electron donors and electron transfer processes, respectively. –log10(FDR) refers to the inverse, log-transformed FDR-adjusted enrichment p-value. TF ratio refers to the number of mapped input genes in relation to the total TF’s target genes. Gene and protein coverage refer to the number of mapped input elements in relation to all pathway-mapped elements. The entire list of significantly enriched metabolic TF and pathways can be found in **Table S3** and **S4**.

### 3.5 Astrocytes and microglia are independently enriched in the AD-metabolic transcriptome

To determine whether cell-type enrichment differences across the AD-metabolic multi-omics datasets could account for the differential pathway over-representation described previously, unconditional EWCE was performed. Significant astrocyte (FDR p-value=0.0000001, standard deviation from the bootstrapped mean or S.D.f.M=7.266) and microglia enrichment (FDR p-value=0.0000001, S.D.f.M=5.770) was found in the AD-metabolic transcriptomics dataset (**Figure 4A**). Oligodendrocyte and astrocyte enrichment in the AD-metabolic proteome lost significance upon multiple-testing correction (FDR p-value=0.07 & S.D.f.M=2.474 and FDR p-value=0.095 & S.D.f.M=2.049 respectively, **Figure 4B**). Astrocyte enrichment was also similarly lost in the GWAS-orthologs dataset (FDR p-value=0.336, S.D.f.M=1.75, **Figure 4C**).

**Figure 4.**
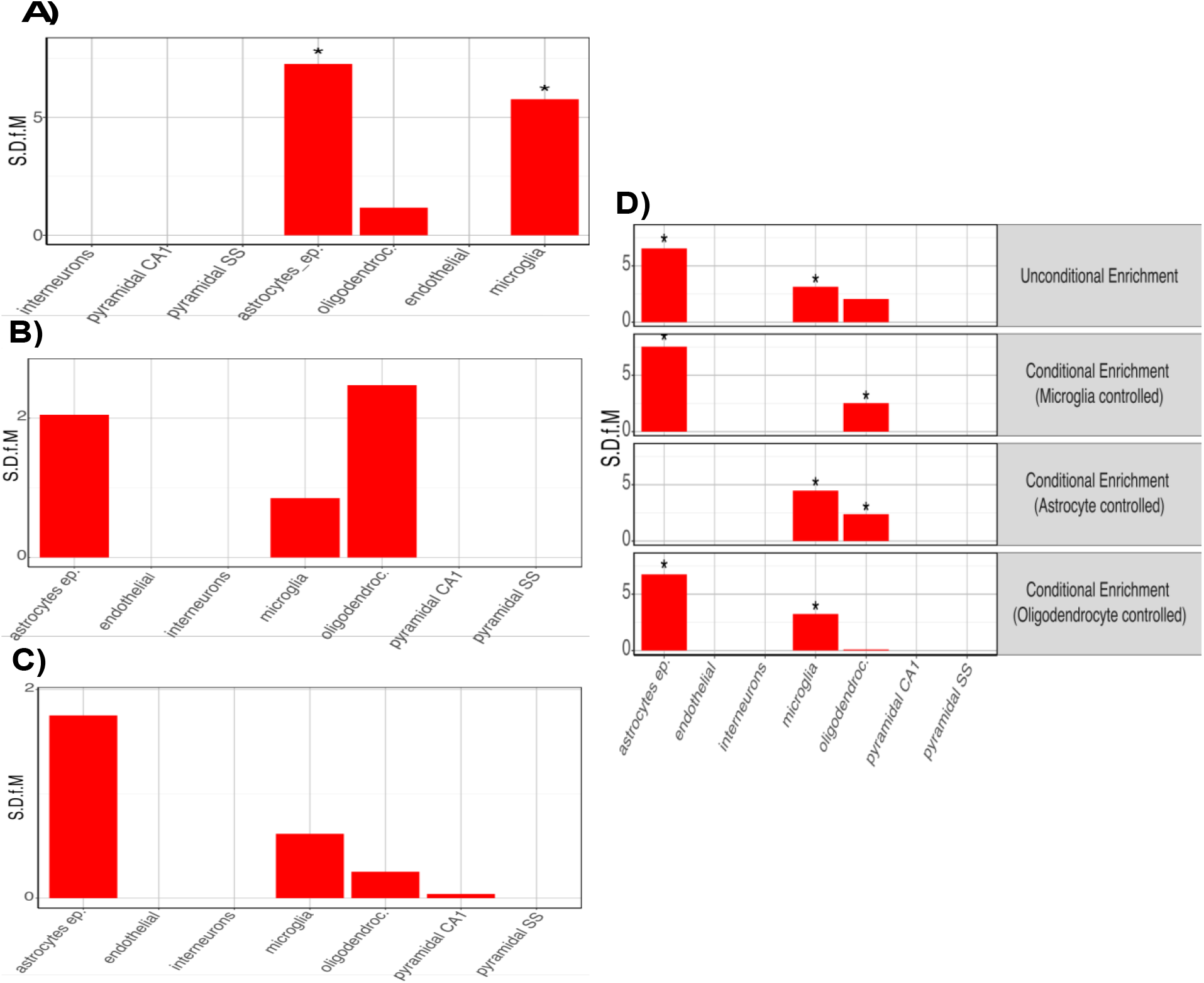
Cell-type enrichment analysis of individual and combined AD-metabolic multi-omics datasets highlight independent astrocyte and microglia enrichment. Unconditional cell type enrichment analysis of AD metabolic (**A**) transcriptomics (**B**) proteomics and (**C**) GWAS-orthologs datasets. (**D**) Conditional cell-type enrichment analysis of combined AD multi-omics dataset. “S.S.f.M” indicates standard deviation from the bootstrapped mean. Asterisk indicates statistical significance upon adjusting for FDR with the Benjamini-Hochberg (B-H) method (p<0.050).

Conditional cell-type enrichment was performed on a combined AD-metabolic multi-omics dataset to investigate enrichment relationships. Controlling for microglia did not ablate astrocytic enrichment (FDR p-value=0.0000001, S.D.f.M=7.540) and vice-versa (FDR p-value=0.0000001, S.D.f.M=4.476), suggesting astrocyte and microglia enrichments were independent of each other (**Figure 4D**). Oligodendrocyte enrichment was however dependent on microglia and astrocytes, as significance was lost upon controlling for either of them (FDR p-value=0.0389 & S.D.f.M=2.531 and FDR p-value=0.0389 & S.D.f.M= 2.387 respectively, **Figure 4D**). Cell-type enrichment statistics can be found in **Table S5**.

### 3.6 Validation of AD multi-omics lipid signatures in ABCA7 KO mice cortex

Given the number of significantly enriched lipid pathways, the results obtained from the multi-omics datasets were validated by comparing them to an internally acquired lipidomics UPLC-MS dataset from cortical extracts of ABCA7-KO and WT mice. To do so, a metabolic subnetwork containing all the significantly enriched lipid pathways was extracted from the generic mouse GSMN (**Figure 3C, Table S4**). This subnetwork involved 119 genes, 81 reactions and 107 metabolites. Of these, 73 were lipid species or terms, as some of them referred to a lipid sub-class, for example a CDP-diacylglycerol, rather than unique species. Upon lipid identifier retrieval, those 73 terms were associated with 133 lipid species, which generated the AD multi-omics predicted lipid signature.

Twenty-eight terms and 60 lipid species from the predicted AD multi-omics lipid signature were found and therefore validated in the ABCA7-KO and WT lipidomes. In particular, 40 lipid species were validated in the negative-mode dataset and 20 species in the positive-mode dataset. The original MS data, containing 5025 and 5811 features in positive and negative ionization mode, respectively, was hence filtered based on these two subsets of lipid species. OPLS-DA was then performed on both original and filtered datasets to assess the presence of any possible separation based on gender and/or genotype, and the potential impact of this feature reduction procedure on the model robustness.

The OPLS-DA model for the negative-mode validated lipid signature was able to separate ABCA7 and WT samples with an even higher degree of robustness than the original dataset (Q^2^cum=0.74 and Q^2^cum=0.56, respectively), which was validated via permutation testing (**Table 2, Figure 5A-B**). As illustrated by the model’s score plot, sample separation was substantially influenced by genotype rather that by variation orthogonal to class membership (**Figure 5B**).

**Table 2.**
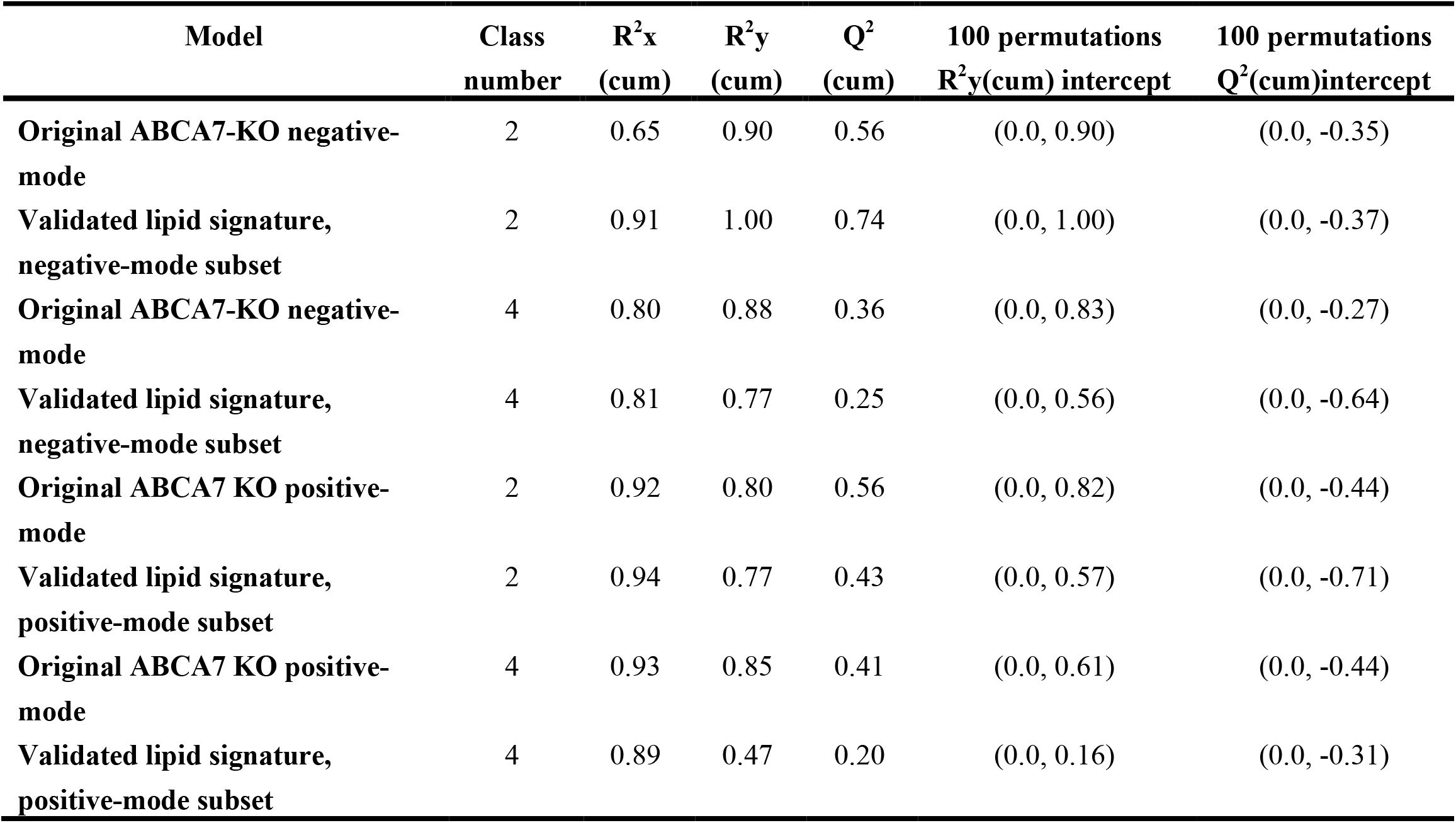
OPLS-DA model parameters for each original ABCA7 dataset and the validated multi-omics lipid signatures subsets.

| Model | Class number | $R^2x$ (cum) | $R^2y$ (cum) | $Q^2$ (cum) | 100 permutations $R^2y(cum)$ intercept | 100 permutations $Q^2(cum)$intercept |
| --- | --- | --- | --- | --- | --- | --- |
| Original ABCA7-KO negative-mode | 2 | 0.65 | 0.90 | 0.56 | (0.0, 0.90) | (0.0, -0.35) |
| Validated lipid signature, negative-mode subset | 2 | 0.91 | 1.00 | 0.74 | (0.0, 1.00) | (0.0, -0.37) |
| Original ABCA7-KO negative-mode | 4 | 0.80 | 0.88 | 0.36 | (0.0, 0.83) | (0.0, -0.27) |
| Validated lipid signature, negative-mode subset | 4 | 0.81 | 0.77 | 0.25 | (0.0, 0.56) | (0.0, -0.64) |
| Original ABCA7 KO positive-mode | 2 | 0.92 | 0.80 | 0.56 | (0.0, 0.82) | (0.0, -0.44) |
| Validated lipid signature, positive-mode subset | 2 | 0.94 | 0.77 | 0.43 | (0.0, 0.57) | (0.0, -0.71) |
| Original ABCA7 KO positive-mode | 4 | 0.93 | 0.85 | 0.41 | (0.0, 0.61) | (0.0, -0.44) |
| Validated lipid signature, positive-mode subset | 4 | 0.89 | 0.47 | 0.20 | (0.0, 0.16) | (0.0, -0.31) |

**Figure 5.**
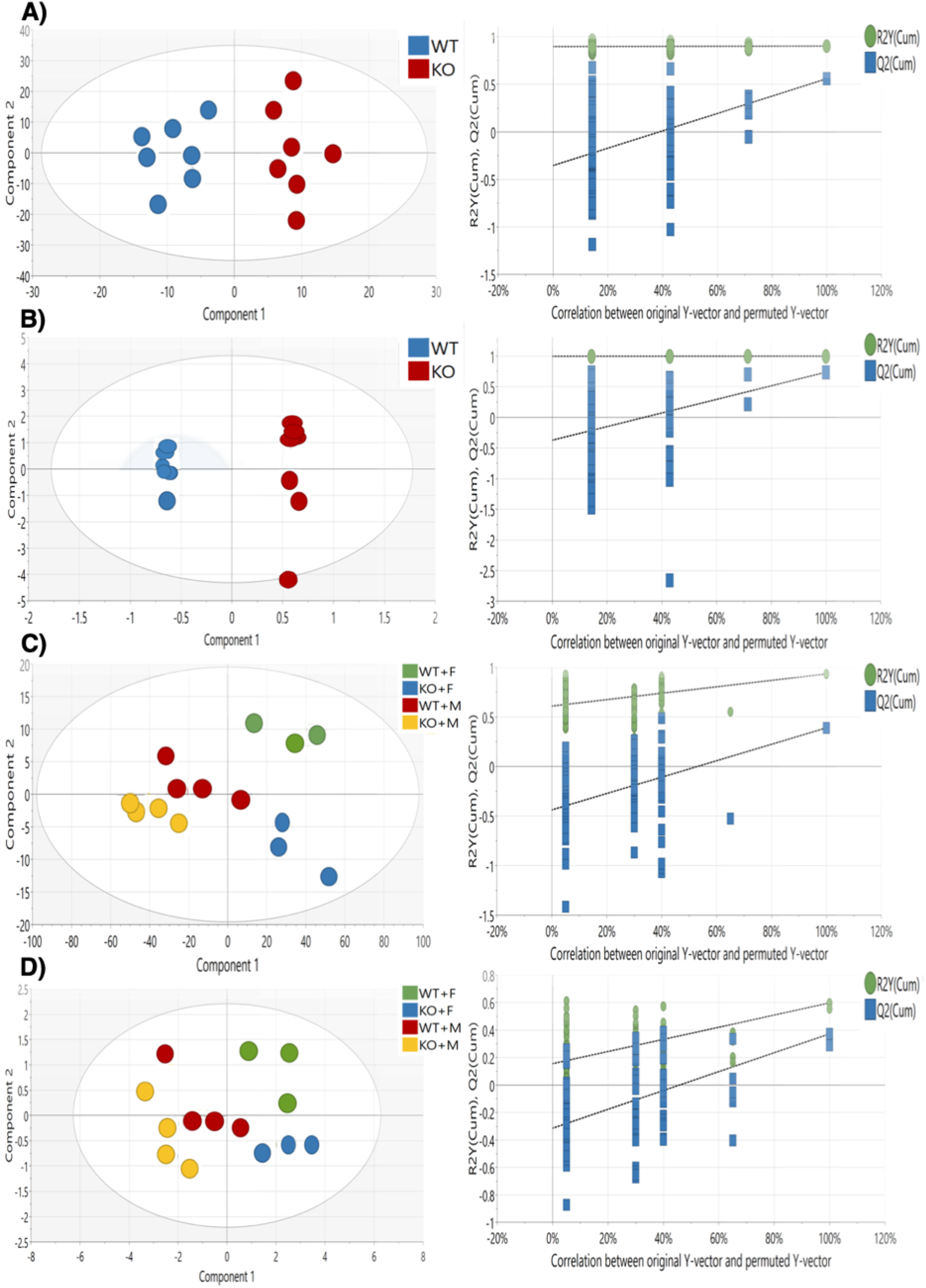
OPLS-DA analysis and permutation test of lipidomics analysis of ABCA7-KO cortical samples and validated lipid signature subsets. OPLS-DA score plot and subsequent 100 permutation test of **A)** Original ABCA7-KO lipidomics dataset in negative mode (R^2^x cum = 0.65, Q^2^ cum = 0.56, R^2^x cum intercept at 0.0, 0.90 and Q^2^ cum intercept at 0.0, -0.35). **B)** ABCA7 KO lipidomics subset corresponding to the validated lipid signature in negative mode (R^2^x cum = 0.91, Q^2^ cum = 0.74, R^2^x cum intercept at 0.0, 1.00 and Q^2^ cum intercept at 0.0, -0.37). **C)** Original ABCA7-KO lipidomics dataset in positive mode (R ^2^x cum = 0.93, Q^2^ cum = 0.41, R^2^x cum intercept at 0.0, 0.61 and Q^2^ cum intercept at 0.0, -0.44). **D)** ABCA7-KO lipidomics subset corresponding to the validated lipid signature in positive mode (R^2^x cum = 0.89, Q^2^ cum = 0.20, R^2^x cum intercept at 0.0, 0.16 and Q^2^ cum intercept at 0.0, -0.31).

Genotype separation was also captured in the OPLS-DA models for the positive-mode original dataset, although less readily differentiated than its negative-mode counterpart (**Table 2**). The robustness of the OPLS-DA model assessing genotype separation for the positive-mode validated lipid signature was impacted by the presence of an outlier (**Table 2**). A strong genotype-sex interaction influenced sample separation in the original positive-mode cortical dataset (Q^2^cum=0.406, **Figure 5C**), but not in the negative mode cortical dataset (Q^2^cum=0.36, **Table 2**). Since the AD multi-omics datasets did not consider sex composition, the positive-mode validated lipid signature should not account for genotype-sex interactions either. Indeed, the genotype-sex interaction was not recapitulated in the positive–mode validated signature subset (Q ^2^cum=0.20, **Table 2, Figure 5D**), while the same model for the negative subset was not calculated due to the lack of statistical power on the correspondent analysis of the original dataset. Therefore, the validated lipid signature in the negative mode seemed robustly influenced by ABCA7 genotype.

We then inspected the VIP scores of the original ABCA7 datasets to investigate whether the predicted lipid signature could play a role in driving class separation in relation to the entire ABCA7 lipidome. Out of the 17 predicted lipid species with a VIP score above 1 in the original ABCA7 lipidome (**Table 3**), 11 were significantly modulated, suggesting the AD multi-omics lipid signature was able to successfully predict significant changes in the ABCA7 cortical lipidome.

**Table 3.** 17 predicted lipid species in the AD multi-omics datasets with a VIP score > 1 in the ABCA7 cortical lipidome.

| Predicted lipid species | GSMN's ID | Detected lipids | LIPIDMAPS ID | Ionization mode | VIP score | Statistical test |
| --- | --- | --- | --- | --- | --- | --- |
| A fatty aldehyde | Fatty-Aldehydes | $C_{26}H_{52}O$ | LMFA06000107 | Negative | 1.45 | 0.455 |
| A saturated-Fatty-AcylCoA | Saturated Fatty-acyl CoA | $C_{40}H_{72}N_7O_{18}P_3S$ | LMFA07050225 | Negative | 1.32 | 0.0530 |
| Lathosterol | CPD-4186 | $C_{27}H_{46}O$ | LMST01010089 | Negative | 1.19 | 0.0070* |
| A L-1-phosphatidyl-glycerol | L-1-PHOSPHA TIDYL-GLYCEROL | $C_{49}H_{85}O_{10}P$ | LMGP04010004 | Negative | 1.14 | 0.1282 |
| A Phosphatidyl-choline | PHOSPHATIDYL CHOLINE | $C_{44}H_{88}NO_8P$ | LMGP01010006 | Negative | 1.13 | 0.6200 |
| Cholesterol | CHOLESTEROL | $C_{27}H_{46}O$ | LMST01010001 | Negative | 1.11 | 0.0262* |
| A fatty acid | Fatty-Acids | $C_{22}H_{37}NO_2$ | LMFA08040001 | Negative | 1.04 | 0.5350 |
| | | $C_{18}H_{30}O_2$ | LMFA01030152 | Positive | 1.09 | 0.0268\$ |
| A 1-acyl glycerophosphocholines | 1-Acylglycero-Phosphocholines | $C_{28}H_{50}NO_7P$ | LMGP01050140 | Negative & Positive | 1.03 | 0.5530 |
| | | $C_{26}H_{52}NO_7P$ | LMGP01050138 | Positive | 1.17 | 0.0342\$ |
| A CDP-diacyl-glycerol | CDPDIACYL-GLYCEROL | $C_{48}H_{85}N_3O_{15}P_2$ | LMGP13010004 | Negative | 1.09 | 0.0273* |
| A diacylglycerol | DIACYL GLYCEROL | $C_{35}H_{68}O_5$ | LMGL02010001 | Positive | 1.50 | 0.0291\$ |
| | | $C_{39}H_{76}O_5$ | LMGL02010002 | Positive | 1.18 | 0.0227\$ |
| 4α-hydroxymethyl-<br>4β-methyl-5α-<br>cholesta-8,24-dien-<br>3β-ol | CPD-4575 | C<sub>29</sub>H<sub>48</sub>O<sub>2</sub> | LMST01010232 | Positive | 1.44 | 0.0291<sup>\$</sup> |
| Ubiquinol-8 | CPD-9956 | C<sub>49</sub>H<sub>74</sub>O<sub>4</sub> | LMPR02010005 | Positive | 1.07 | 0.0247<sup>\$</sup> |
| An acyl-sn-Glycerol-3phosphate | ACYL-SN-GLYCEROL-3P | C<sub>21</sub>H<sub>43</sub>O<sub>7</sub>P | LMGP10050005 | Positive | 1.07 | 0.0295<sup>\$</sup> |
| 7-dehydro-cholesterol | CPD-4187 | C<sub>27</sub>H<sub>44</sub>O | LMST01010069 | Positive | 1.04 | 0.0427<sup>\$</sup> |

Predicted lipid signature was derived from an extracted metabolic subnetwork containing all significantly enriched lipid metabolic pathways in the AD transcriptomics and proteomics datasets, which contained 73 lipid terms. If species in the predicted lipid signature referred to a lipid class, all of the detected compounds belonging to that lipid class were considered for the analysis. This approach yielded 133 unique lipid species, which were mapped to 60 and 20 lipids detected in negative and positive ion mode, respectively. Of these predicted lipid species, 17 had a VIP score > 1 in the OPLS - DA models for the original ABCA7 datasets. *References to p <0.050 significance upon unpaired t-test and Mann Whitney non-parametric testing on intensity differences between ABCA7 and WT mice in the original negative mode ABCA7 dataset. ^$^ refers to significance upon One-way ANOVA using B-H correction for multiple testing on differences between ABCA7-males and ABCA7-females, WT-females or WT-males in the original positive mode ABCA7 dataset.

### 3.7 Validation of lipid-AD risk loci associations in the Airwave and RS cohorts

Lastly, we performed a MWAS using ^1^H NMR spectra of human blood serum from 3258 individuals from the Airwave and RS cohorts (Elliott *et al*. 2014; Ikram *et al*. 2020). As these cohorts consist of predominantly healthy individuals, we used 47 known AD risk loci to identify AD risk carriers (Lambert *et al*. 2013; Jansen *et al*. 2019; Kunkle *et al*. 2019).

After performing MWAS, we detected 298 SNP-metabolite associations from the three NMR pulse sequences, out of which 107 in the lipoprotein, 13 in the CPMG, and 178 in the solvent suppression pulse sequence spectra datasets. Association with *APOE* was found for 83% of these, reflecting the importance of this gene in regulating components of the blood metabolome (**Figure 6a**).

**Figure 6:**
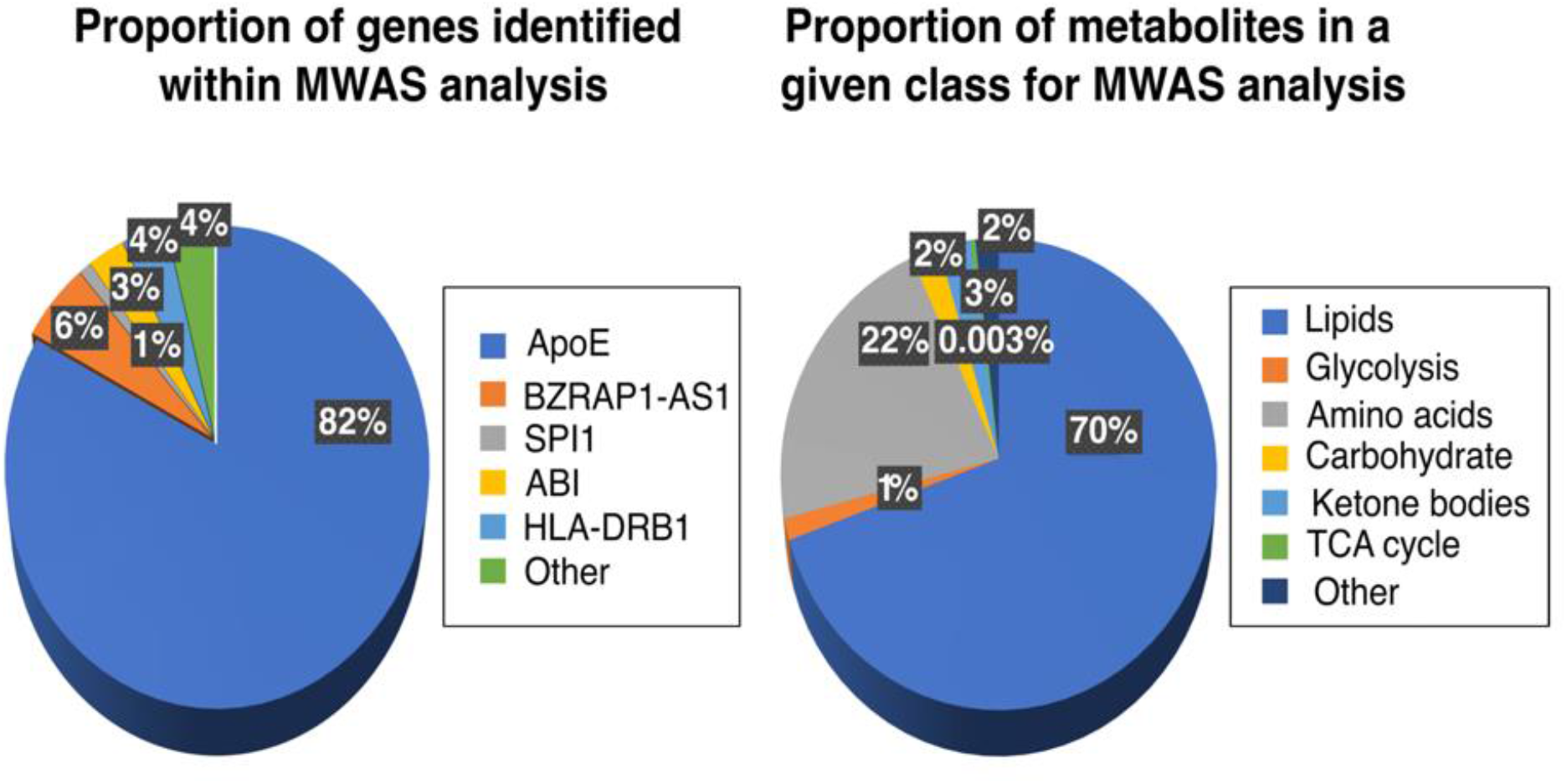
Metabolome-wide association study of the blood metabolome for AD risk genes in the Airwave and RS cohorts. **A)** Proportion of AD risk genes significantly associated with fluctuating metabolite levels detected in the blood samples of individuals in the Airwave and RS cohorts. MWSL was set to 0.05 upon 10,000 permutations to control for FWER. **B)** Proportion of metabolite classes associated with AD risk loci in the Airwave and RS cohorts. MWSL was set to 0.05 upon 10,000 permutations to control for FWER.

To examine the associations further we classified the detected metabolites according to their chemical characteristics and biological role into lipids, amino acids, carbohydrates, glycolysis intermediates, TCA cycle intermediates, ketone bodies and other metabolites. Lipids included resonances that were associated with both protons attached to the fatty acid and the head group (largely choline and glycerol) along with protons from cholesterol and cholesterol esters. The dominant class was represented by lipids, comprising over 70% of the associations (**Figure 6b**).

## 4 Discussion

The main aim of this study was to validate the presence of metabolic perturbations in AD using multi-omics pathway-based integration and metabolic-subnetwork extraction. We hypothesized that metabolic alterations detected at multiple omics levels could predict a robust metabolic signature in the AD metabolome. If validated, these results would provide a comprehensive perspective on AD metabolism while supporting the use of GSMNs to identify consistent metabolic alterations in AD.

GO analysis of AD transcriptomics, proteomics and GWAS-orthologs data revealed numerous enriched metabolic BP. Although the initial mapping of DE transcripts, proteins and GWAS-orthologs certainly removed elements with no metabolic roles, this step did not disproportionately influence metabolic BP term over-representation *per se*, as only 3 out of 11 BP in mapped GWAS-orthologs were metabolic. Lipid and fatty-acid BP enrichment was found across the AD all-mapped transcriptome and proteome. This observation was further supported by *TAL1, MAX* and *BHLHE40* over-representation in both datasets. *TAL1* modulates lipid metabolism in the context of cell membrane integrity (Kassouf *et al*. 2010), *MAX-MYC* interaction strongly dysregulates fatty-acid metabolism in neurodegeneration (Carroll *et al*. 2018) and *BHLHE40* is necessary for insulin-mediated *SREBP1* induction, a lipid homeostasis regulator (Tian *et al*. 2018).

Pathway and TF enrichment analysis implicated differential metabolic processes across the AD multi-omics datasets, which also exhibited different cell-type enrichments. Cholesterol biosynthesis, phospholipases, fatty-acid metabolism and *SREBF1* were strongly enriched in the AD-metabolic transcriptome, which also exhibited astrocyte and microglia cell-type enrichment. These multi-level results provide further evidence supporting the existence of wide-spread lipidomic alterations in AD microglia (Wang *et al*. 2015). Previously, an allele variant in the *SREPF1* gene was found to be neuroprotective in APOE4 carriers in terms of dementia incidence (Spell *et al*. 2004). Extensive lipidome changes are present in TREM2 - defficent microglia, another gene variant heavily implicated in AD pathogenesis (Nugent *et al*. 2020). Phospholipase-amyloid interactions seem to facilitate microglia Aβ endocytosis, therefore contributing to neuroinflammation (Teng *et al*. 2019). Aerobic respiration, TCA cycle and glycolysis were enriched in the AD-metabolic proteome; these pathways are consistent with signs of mitochondrial dysfunction that are commonly found in neurodegeneration (Wang *et al*. 2020). Indeed, significant energy metabolism deficits have been detected in human(Johnson *et al*. 2020) and AD mice brain proteomes (Yu *et al*. 2018).

The main finding in this study is the validation of a predicted lipid signature derived from an extracted metabolic subnetwork with all significantly enriched lipid pathways in AD multi-omics datasets. The OPLS-DA model for the validated lipid signature in negative ion mode LC-MS dataset was capable of driving class separation based on ABCA7 genotype with a higher degree of robustness than in the original dataset; the reduced number of features was not a confounding factor for the model, but instead allowed for the removal of features originally decreasing the model robustness. Multi-omics integration is being increasingly used to draw biologically meaningful conclusions over large datasets (Pinu *et al*. 2019), and has been previously applied to AD data to infer metabolic perturbations using protein ranking and gene-set enrichment (Bundy *et al*. 2019; Bai *et al*. 2020), gene-protein interaction networks (Canchi *et al*. 2019) and protein-protein interaction networks (Zhang *et al*. 2020). To our knowledge, this is the first study using multi-omics pathway-based integration and metabolic subnetwork extraction to identify and subsequently validate a lipid metabolic signature in the AD lipidome.

Eleven lipid species from the validated lipid signature were significantly modulated in the cortical ABCA7 lipidome, of which four belonged to the cholesterol biosynthesis pathway. Lathosterol and cholesterol were significantly decreased in the ABCA7-KO lipidome compared to WT, whereas 7-dehydro-cholesterol and 4α-hydroxymethyl-4β-methyl-5α-cholesta-8,24-dien-3β-ol were significantly decreased in ABCA7-females compared to ABCA7-males. The evidence is mixed regarding cholesterol and intermediate sterols changes in ABCA7 mice. One study showed no cholesterol changes in ABCA7-KO mice brains (Satoh *et al*. 2015); serum cholesterol levels were however decreased in female ABCA7-KO mice (Kim *et al*. 2005). This study appears more aligned with the latter, as decreased free - cholesterol levels and sex-specific sterol intermediates differences were detected. This discrepancy is extended to other AD mouse models. Free-cholesterol and lathosterol levels exhibited non-significant changes in TgCRND8 (Yang *et al*. 2014) and APP/PS1 mice (Bogie *et al*. 2019), whereas lanosterol and cholesteryl acetate were up-regulated in APOE4 mice (Nuriel *et al*. 2017). Despite these disagreements, the importance of sterol intermediates in AD is reflected therapeutically, as a recent drug-repurposing screen identified several tau - reducing compounds which targeted cholesterol-esters (van der Kant *et al*. 2019).

We also performed an MWAS analysis using SNPs previously associated with LOAD and metabolites detected in blood plasma from the Airwave and Rotterdam cohorts using ^1^H NMR spectroscopy. Mean ages of recruitment in these cohorts are relatively young, and thus our reported 298 SNP-metabolite associations may represent early stages of the disease, as the brain begins to accumulate neurodegenerative features that ultimately results in Mild Cognitive Impairment (MCI) and AD. Using three distinct NMR pulse sequences, we were able to detect a range of metabolites including lipids, amino acids, glycolysis, TCA cycle intermediates and ketone bodies. Lipids were the commonest metabolite class represented in metabolite-SNP associations, suggesting that dysregulation of lipid metabolism may be some of the earliest events in AD.

There are important limitations associated with this study. Firstly, this study included multi-omics data from several brain regions, ages and AD mouse models. Therefore, region and age-specific TF upstream-regulation and metabolic alterations that are frequent in AD (González-Domínguez *et al*. 2014) were not assessed. It is also notoriously difficult to annotate lipid species into GSMNs due to the complexities associated with lipid nomenclature and identification (Poupin *et al*. 2020). This study successfully overcame this limitation by allowing second-order lipid species matching to their associated broader lipid term whenever unique lipid species matching was not possible (Poupin *et al*. 2020). This study was also limited in that cell type enrichment analysis could not distinguish whether astrocytic and microglia enrichment was associated with gliosis in disease rather than AD pathology *per se*, as cell type proportions could not be adequately controlled *in silico*. Additionally, APOE-associated SNPs dominated our MWAS analysis, which could be attributed to the known association of ApoE with dyslipidemia and atherosclerosis (Bouchareychas & Raffai 2018). Furthermore, ^1^H NMR spectra of blood plasma detect a high proportion of lipids compared with other classes of metabolites and is relatively insensitive as a technique. We are currently performing mass spectrometry to expand the coverage of the metabolome to further investigate the earliest molecular events in AD.

## 5 Conclusions

In summary, this study highlights the suitability of integrating multi-omics data into GSMNs to identify metabolic alterations in AD. Pathway-based integration of multi-omics data revealed distinct perturbations in lipid metabolism in the AD mouse brain. Predicted lipids extracted from the over-represented lipid pathway’s metabolic subnetwork was validated in the ABCA7 lipidome, with its associated multi-variate model robustly modelling class separation. Furthermore, more than 70% of 298 SNP-metabolite associations in a MWAS corresponded to lipid species, thus validating the presence of lipidomic dysregulation in AD.

## Data Availability

We reference all data used on mouse studies which is freely available through omics repositories. These links are provided in the text.
In addition, ethical approval for access to the Airwave cohort was granted following application to the access committee via the Dementia Platform UK (https://portal.dementiasplatform.uk/). Access to the RS cohort was granted following access to the Management Committee and conducted under approval from the Ministry of Health, Welfare and Sport of the Netherlands.

## Abbreviations

Airwave: Airwave Health Monitoring Study
AD: Alzheimer’s Disease
Aβ: amyloid-beta
APP: amyloid-precursor protein
ABCA7: ATP-binding-cassette subfamily-A member-7 gene
APOE: apolipoprotein epsilon
ChEA3: ChIP-X enrichment analysis 3
DAVID: database for annotation, visualization and integrated discovery
DE: differentially expressed
EWCE: expression weighted cell-type enrichment
FDR: false discovery rate
FC: fold change
GEO: gene expression omnibus
GRCh37: genome reference consortium-human build-37
GSMN: genome-scale metabolic networks
GWAS: genome-wide association studies
IGAP: international genomics of Alzheimer’s cohorts
iTRAQ: isobaric tag for relative and absolute quantification
KO: knock-out
MAGMA: multi-marker analysis of genomic annotation
MWAS: metabolome-wide association study
NMR: nuclear magnetic resonance
OPLS-DA: orthogonal projections to latent structures-discriminant analysis
PQN: probabilistic quotient normalization
PRIDE: protein identification database
RP-UPLC-MS: reverse-phase ultraperformance liquid chromatography-mass spectrometry
RS: Rotterdam study
SAM: significance analysis of microarray
SNPs: single nucleotide polymorphisms
S.D.f.M: standard deviation from the bootstrapped mean
SREBP2: sterol regulatory element binding protein 2
TF: transcription factor
TREM2: triggering receptor expressed on myeloid cells-2
UPLC-MS: ultraperformance liquid chromatography-mass spectrometry
VIP: variable influence of projection
WT: wild-type

## Author Contributions

M.E.G.S. and J.L.G. conceived and designed the study. M.E.G.S. retrieved and analyzed the transcriptomics, proteomics, GWAS and lipidomics data. B.R.D. acquired the lipidomic data. S.L. and B.R.D. processed the lipidomic data. I.K. performed the MWAS study, which used data from two on-going cohorts oversaw by P.E. M.E.G.S. and J.L.G. interpreted the data. M.E.G.S. drafted the manuscript, which received critical input from J.L.G. All authors have read and approved the published version of the manuscript.

## Acknowledgments

The authors would like to acknowledge Dr. Tomonori Aikawa and Professor Takahisa Kanekiyo from the Mayo Clinic, Jacksonville, Florida for providing the ABCA7 cortical mouse tissue.

## Supplementary Materials

The following are available : **Table S1**: Transcription Factor enrichment analysis of all-mapped AD transcriptomics and proteomics datasets; **Table S2**: Biological Process (BP), Molecular Function (MF) and Cellular Compartment (CC) enrichment analysis of all-mapped AD transcriptomics, proteomics and GWAS-orthologs datasets; **Table S3**: Transcription Factor enrichment analysis of AD-metabolic transcriptomics and proteomics datasets; **Table S4**: Metabolic pathway enrichment analysis of AD-metabolic transcriptomics, proteomics and GWAS-orthologs datasets; **Table S5**: Unconditional and conditional EWCE analysis of AD-metabolic multi-omics datasets.

## References

Aikawa, T., Holm, M. L. and Kanekiyo, T. (2018) ABCA7 and Pathogenic Pathways of Alzheimer’s Disease. Brain Sci 8.

Andreas, N. J., Basu Roy, R., Gomez-Romero, M. et al. (2020) Performance of metabonomic serum analysis for diagnostics in paediatric tuberculosis. Sci Rep 10, 7302.

Auton, A., Brooks, L. D., Durbin, R. M. et al. (2015) A global reference for human genetic variation. Nature 526, 68–74.

Aydin, D., Filippov, M. A., Tschäpe, J. A., Gretz, N., Prinz, M., Eils, R., Brors, B. and Müller, U. C. (2011) Comparative transcriptome profiling of amyloid precursor protein family members in the adult cortex. BMC Genomics 12, 160.

Bai, B., Wang, X., Li, Y. et al. (2020) Deep Multilayer Brain Proteomics Identifies Molecular Networks in Alzheimer’s Disease Progression. Neuron 105, 975–991.e977.

Bogie, J., Hoeks, C., Schepers, M. et al. (2019) Dietary Sargassum fusiforme improves memory and reduces amyloid plaque load in an Alzheimer’s disease mouse model. Sci Rep 9, 4908.

Bouchareychas, L. and Raffai, R. L. (2018) Apolipoprotein E and Atherosclerosis: From Lipoprotein Metabolism to MicroRNA Control of Inflammation. J Cardiovasc Dev Dis 5.

Bundy, J. L., Vied, C., Badger, C. and Nowakowski, R. S. (2019) Sex-biased hippocampal pathology in the 5XFAD mouse model of Alzheimer’s disease: A multi-omic analysis. J Comp Neurol 527, 462–475.

Buniello, A., MacArthur, J. A. L., Cerezo, M. et al. (2019) The NHGRI-EBI GWAS Catalog of published genome-wide association studies, targeted arrays and summary statistics 2019. Nucleic Acids Res 47, D1005–d1012.

Canchi, S., Raao, B., Masliah, D., Rosenthal, S. B., Sasik, R., Fisch, K. M., De Jager, P. L., Bennett, D. A. and Rissman, R. A. (2019) Integrating Gene and Protein Expression Reveals Perturbed Functional Networks in Alzheimer’s Disease. Cell Rep 28, 1103–1116.e1104.

Canzler, S., Schor, J., Busch, W. et al. (2020) Prospects and challenges of multi-omics data integration in toxicology. Arch Toxicol 94, 371–388.

Carroll, P. A., Freie, B. W., Mathsyaraja, H. and Eisenman, R. N. (2018) The MYC transcription factor network: balancing metabolism, proliferation and oncogenesis. Front Med 12, 412–425.

Caspi, R., Billington, R., Ferrer, L. et al. (2016) The MetaCyc database of metabolic pathways and enzymes and the BioCyc collection of pathway/genome databases. Nucleic Acids Res 44, D471–480.

Castagné R, Boulangé CL, Karaman I et al. (2017) Improving Visualization and Interpretation of Metabolome-Wide Association Studies: An Application in a Population-Based Cohort Using Untargeted (1)H NMR Metabolic Profiling. J Proteome Res 16, 3623–3633.

Chadeau-Hyam M, Ebbels TM, Brown IJ et al. (2010) Metabolic profiling and the metabolome-wide association study: significance level for biomarker identification. J Proteome Res. 9, :4620–4627.

Chazalviel, M., Frainay, C., Poupin, N., Vinson, F., Merlet, B., Gloaguen, Y., Cottret, L. and Jourdan, F. (2018) MetExploreViz: web component for interactive metabolic network visualization. Bioinformatics 34, 312–313.

Clough, E. and Barrett, T. (2016) The Gene Expression Omnibus Database. Methods Mol Biol 1418, 93–110.

Cottret, L., Frainay, C., Chazalviel, M. et al. (2018) MetExplore: collaborative edition and exploration of metabolic networks. Nucleic Acids Res 46, W495–w502.

Cutler, R. G., Kelly, J., Storie, K., Pedersen, W. A., Tammara, A., Hatanpaa, K., Troncoso, J. C. and Mattson, M. P. (2004) Involvement of oxidative stress-induced abnormalities in ceramide and cholesterol metabolism in brain aging and Alzheimer’s disease. Proc Natl Acad Sci U S A 101, 2070–2075.

Davis, C. A., Hitz, B. C., Sloan, C. A. et al. (2018) The Encyclopedia of DNA elements (ENCODE): data portal update. Nucleic Acids Res 46, D794–d801.

Davis, S. and Meltzer, P. S. (2007) GEOquery: a bridge between the Gene Expression Omnibus (GEO) and BioConductor. Bioinformatics 23, 1846–1847.

de Leeuw, C. A., Mooij, J. M., Heskes, T. and Posthuma, D. (2015) MAGMA: generalized gene-set analysis of GWAS data. PLoS Comput Biol 11, e1004219.

De Roeck, A., Van Broeckhoven, C. and Sleegers, K. (2019) The role of ABCA7 in Alzheimer’s disease: evidence from genomics, transcriptomics and methylomics. Acta Neuropathol 138, 201–220.

Dennis, G. Jr., Sherman, B. T., Hosack, D. A., Yang, J., Gao, W., Lane, H. C. and Lempicki, R. A. (2003) DAVID: Database for Annotation, Visualization, and Integrated Discovery. Genome Biol 4, P3.

DeTure, M. A. and Dickson, D. W. (2019) The neuropathological diagnosis of Alzheimer’s disease. Mol Neurodegener 14, 32.

Dona A.C., Jiménez B., Schäfer H. et al. (2014) Precision high-throughput proton NMR spectroscopy of human urine, serum, and plasma for large-scale metabolic phenotyping. Analytical Chemistry 86, 9887–9894.

Elliott, P., Vergnaud, A. C., Singh, D., Neasham, D., Spear, J. and Heard, A. (2014) The Airwave Health Monitoring Study of police officers and staff in Great Britain: rationale, design and methods. Environ Res 134, 280–285.

Fahy, E., Subramaniam, S., Murphy, R. C. et al. (2009) Update of the LIPID MAPS comprehensive classification system for lipids. J Lipid Res 50 Suppl, S9–14.

Faivre, E., Coelho, J. E., Zornbach, K. et al. (2018) Beneficial Effect of a Selective Adenosine A(2A) Receptor Antagonist in the APPswe/PS1dE9 Mouse Model of Alzheimer’s Disease. Front Mol Neurosci 11, 235.

Fang, E. F., Hou, Y., Palikaras, K. et al. (2019) Mitophagy inhibits amyloid-β and tau pathology and reverses cognitive deficits in models of Alzheimer’s disease. Nat Neurosci 22, 401–412.

Fernandez, C. G., Hamby, M. E., McReynolds, M. L. and Ray, W. J. (2019) The Role of APOE4 in Disrupting the Homeostatic Functions of Astrocytes and Microglia in Aging and Alzheimer’s Disease. Front Aging Neurosci 11, 14.

Frainay, C. and Jourdan, F. (2017) Computational methods to identify metabolic sub - networks based on metabolomic profiles. Brief Bioinform 18, 43–56.

González-Domínguez, R., García-Barrera, T., Vitorica, J. and Gómez-Ariza, J. L. (2014) Region-specific metabolic alterations in the brain of the APP/PS1 transgenic mice of Alzheimer’s disease. Biochim Biophys Acta 1842, 2395–2402.

Griffin, J. L., Liggi, S. and Hall, Z. (2020) CHAPTER 2 Multivariate Statistics in Lipidomics. In: Lipidomics: Current and Emerging Techniques, pp. 25–48. The Royal Society of Chemistry.

Hamezah, H. S., Durani, L. W., Yanagisawa, D., Ibrahim, N. F., Aizat, W. M., Makpol, S., Wan Ngah, W. Z., Damanhuri, H. A. and Tooyama, I. (2019) Modulation of Proteome Profile in AβPP/PS1 Mice Hippocampus, Medial Prefrontal Cortex, and Striatum by Palm Oil Derived Tocotrienol-Rich Fraction. J Alzheimers Dis 72, 229–246.

Hamilton, L. K., Dufresne, M., Joppé, S. E. et al. (2015) Aberrant Lipid Metabolism in the Forebrain Niche Suppresses Adult Neural Stem Cell Proliferation in an Animal Model of Alzheimer’s Disease. Cell Stem Cell 17, 397–411.

Hou, Y., Lautrup, S., Cordonnier, S. et al. (2018) NAD(+) supplementation normalizes key Alzheimer’s features and DNA damage responses in a new AD mouse model with introduced DNA repair deficiency. Proc Natl Acad Sci U S A 115, E1876–e1885.

Ikram, M. A., Brusselle, G., Ghanbari, M. et al. (2020) Objectives, design and main findings until 2020 from the Rotterdam Study. Eur J Epidemiol 35, 483–517.

Jansen, I. E., Savage, J. E., Watanabe, K. et al. (2019) Genome-wide meta-analysis identifies new loci and functional pathways influencing Alzheimer’s disease risk. Nat Genet 51, 404–413.

Jiménez, B., Holmes, E., Heude, C. et al. (2018) Quantitative Lipoprotein Subclass and Low Molecular Weight Metabolite Analysis in Human Serum and Plasma by (1)H NMR Spectroscopy in a Multilaboratory Trial. Anal Chem 90, 11962–11971.

Johnson, E. C. B., Dammer, E. B., Duong, D. M. et al. (2020) Large-scale proteomic analysis of Alzheimer’s disease brain and cerebrospinal fluid reveals early changes in energy metabolism associated with microglia and astrocyte activation. Nat Med 26, 769–780.

Jones, P., Côté, R. G., Martens, L., Quinn, A. F., Taylor, C. F., Derache, W., Hermjakob, H. and Apweiler, R. (2006) PRIDE: a public repository of protein and peptide identifications for the proteomics community. Nucleic Acids Res 34, D659–663.

Kassouf, M. T., Hughes, J. R., Taylor, S., McGowan, S. J., Soneji, S., Green, A. L., Vyas, P. and Porcher, C. (2010) Genome-wide identification of TAL1’s functional targets: insights into its mechanisms of action in primary erythroid cells. Genome Res 20, 1064–1083.

Keenan, A. B., Torre, D., Lachmann, A. et al. (2019) ChEA3: transcription factor enrichment analysis by orthogonal omics integration. Nucleic Acids Res 47, W212–w224.

Kim, D. K., Han, D., Park, J. et al. (2019) Deep proteome profiling of the hippocampus in the 5XFAD mouse model reveals biological process alterations and a novel biomarker of Alzheimer’s disease. Exp Mol Med 51, 1–17.

Kim, W. S., Fitzgerald, M. L., Kang, K. et al. (2005) Abca7 null mice retain normal macrophage phosphatidylcholine and cholesterol efflux activity despite alterations in adipose mass and serum cholesterol levels. J Biol Chem 280, 3989–3995.

Kunkle, B. W., Grenier-Boley, B., Sims, R. et al. (2019) Genetic meta-analysis of diagnosed Alzheimer’s disease identifies new risk loci and implicates Aβ, tau, immunity and lipid processing. Nat Genet 51, 414–430.

Lachen-Montes, M., González-Morales, A., Palomino, M. et al. (2019) Early-Onset Molecular Derangements in the Olfactory Bulb of Tg2576 Mice: Novel Insights Into the Stress-Responsive Olfactory Kinase Dynamics in Alzheimer’s Disease. Front Aging Neurosci 11, 141.

Lambert, J. C., Ibrahim-Verbaas, C. A., Harold, D. et al. (2013) Meta-analysis of 74,046 individuals identifies 11 new susceptibility loci for Alzheimer’s disease. Nat Genet 45, 1452–1458.

Liggi, S., Hinz, C., Hall, Z., Santoru, M. L., Poddighe, S., Fjeldsted, J., Atzori, L. and Griffin, J. L. (2018) KniMet: a pipeline for the processing of chromatography-mass spectrometry metabolomics data. Metabolomics 14, 52.

Liu, K. D., Acharjee, A., Hinz, C. et al. (2020) Consequences of Lipid Remodeling of Adipocyte Membranes Being Functionally Distinct from Lipid Storage in Obesity. J Proteome Res 19, 3919–3935.

Mancuso, R., Van Den Daele, J., Fattorelli, N. et al. (2019) Stem-cell-derived human microglia transplanted in mouse brain to study human disease. Nat Neurosci 22, 2111–2116.

Marioni, R. E., Harris, S. E., Zhang, Q. et al. (2018) GWAS on family history of Alzheimer’s disease. Transl Psychiatry 8, 99.

Marsh, S. E., Abud, E. M., Lakatos, A. et al. (2016) The adaptive immune system restrains Alzheimer’s disease pathogenesis by modulating microglial function. Proc Natl Acad Sci U S A 113, E1316–1325.

Murgia, F., Muroni, A., Puligheddu, M. et al. (2017) Metabolomics As a Tool for the Characterization of Drug-Resistant Epilepsy. Front Neurol 8, 459.

Nugent, A. A., Lin, K., van Lengerich, B. et al. (2020) TREM2 Regulates Microglial Cholesterol Metabolism upon Chronic Phagocytic Challenge. Neuron 105, 837–854.e839.

Nuriel, T., Angulo, S. L., Khan, U. et al. (2017) Neuronal hyperactivity due to loss of inhibitory tone in APOE4 mice lacking Alzheimer’s disease-like pathology. Nat Commun 8, 1464.

Oberg, A. L. and Mahoney, D. W. (2012) Statistical methods for quantitative mass spectrometry proteomic experiments with labeling. BMC Bioinformatics 13 Suppl 16, S7.

Palomino-Alonso, M., Lachén-Montes, M., González-Morales, A., Ausín, K., Pérez-Mediavilla, A., Fernández-Irigoyen, J. and Santamaría, E. (2017) Network-Driven Proteogenomics Unveils an Aging-Related Imbalance in the Olfactory IκBα-NFκB p65 Complex Functionality in Tg2576 Alzheimer’s Disease Mouse Model. Int J Mol Sci 18.

Pappireddi, N., Martin, L. and Wühr, M. (2019) A Review on Quantitative Multiplexed Proteomics. Chembiochem 20, 1210–1224.

Perez Ortiz, J. M. and Swerdlow, R. H. (2019) Mitochondrial dysfunction in Alzheimer’s disease: Role in pathogenesis and novel therapeutic opportunities. Br J Pharmacol 176, 3489–3507.

Pinu, F. R., Beale, D. J., Paten, A. M., Kouremenos, K., Swarup, S., Schirra, H. J. and Wishart, D. (2019) Systems Biology and Multi-Omics Integration: Viewpoints from the Metabolomics Research Community. Metabolites 9.

Polito, V. A., Li, H., Martini-Stoica, H. et al. (2014) Selective clearance of aberrant tau proteins and rescue of neurotoxicity by transcription factor EB. EMBO Mol Med 6, 1142–1160.

Poupin, N., Vinson, F., Moreau, A. et al. (2020) Improving lipid mapping in Genome Scale Metabolic Networks using ontologies. Metabolomics 16, 44.

Preuss, C., Pandey, R., Piazza, E. et al. (2020) A novel systems biology approach to evaluate mouse models of late-onset Alzheimer’s disease. Mol Neurodegener 15, 67.

Robinson, O., Chadeau Hyam, M., Karaman, I. et al. (2020) Determinants of accelerated metabolomic and epigenetic aging in a UK cohort. Aging Cell 19, e13149.

Satoh, K., Abe-Dohmae, S., Yokoyama, S., St George-Hyslop, P. and Fraser, P. E. (2015) ATP-binding cassette transporter A7 (ABCA7) loss of function alters Alzheimer amyloid processing. J Biol Chem 290, 24152–24165.

Skene, N. G., Bryois, J., Bakken, T. E. et al. (2018) Genetic identification of brain cell types underlying schizophrenia. Nat Genet 50, 825–833.

Skene, N. G. and Grant, S. G. (2016) Identification of Vulnerable Cell Types in Major Brain Disorders Using Single Cell Transcriptomes and Expression Weighted Cell Type Enrichment. Front Neurosci 10, 16.

Spell, C., Kölsch, H., Lütjohann, D. et al. (2004) SREBP-1a polymorphism influences the risk of Alzheimer’s disease in carriers of the ApoE4 allele. Dement Geriatr Cogn Disord 18, 245–249.

Su, M., Subbaraj, A. K., Fraser, K. et al. (2019) Lipidomics of Brain Tissues in Rats Fed Human Milk from Chinese Mothers or Commercial Infant Formula. Metabolites 9.

Tautenhahn, R., Patti, G. J., Rinehart, D. and Siuzdak, G. (2012) XCMS Online: a web-based platform to process untargeted metabolomic data. Anal Chem 84, 5035–5039.

Team, R. C. (2020) R: A language and environment for statistical computing. R Foundation for Statistical Computing, Vienna, Austria.

Teng, T., Dong, L., Ridgley, D. M., Ghura, S., Tobin, M. K., Sun, G. Y., LaDu, M. J. and Lee, J. C. (2019) Cytosolic Phospholipase A(2) Facilitates Oligomeric Amyloid-β Peptide Association with Microglia via Regulation of Membrane-Cytoskeleton Connectivity. Mol Neurobiol 56, 3222–3234.

Tian, J., Wu, J., Chen, X., Guo, T., Chen, Z. J., Goldstein, J. L. and Brown, M. S. (2018) BHLHE40, a third transcription factor required for insulin induction of SREBP-1c mRNA in rodent liver. Elife 7.

Tusher, V. G., Tibshirani, R. and Chu, G. (2001) Significance analysis of microarrays applied to the ionizing radiation response. Proc Natl Acad Sci U S A 98, 5116–5121.

Tyanova, S., Temu, T., Sinitcyn, P., Carlson, A., Hein, M. Y., Geiger, T., Mann, M. and Cox, J. (2016) The Perseus computational platform for comprehensive analysis of (prote)omics data. Nat Methods 13, 731–740.

Tzoulaki, I., Castagné, R., Boulangé, C. L. et al. (2019) Serum metabolic signatures of coronary and carotid atherosclerosis and subsequent cardiovascular disease. Eur Heart J 40, 2883–2896.

van der Kant, R., Langness, V. F., Herrera, C. M. et al. (2019) Cholesterol Metabolism Is a Druggable Axis that Independently Regulates Tau and Amyloid-β in iPSC-Derived Alzheimer’s Disease Neurons. Cell Stem Cell 24, 363–375.e369.

Wang, E., Zhu, H., Wang, X., Gower, A. C., Wallack, M., Blusztajn, J. K., Kowall, N. and Qiu, W. Q. (2017) Amylin Treatment Reduces Neuroinflammation and Ameliorates Abnormal Patterns of Gene Expression in the Cerebral Cortex of an Alzheimer’s Disease Mouse Model. J Alzheimers Dis 56, 47–61.

Wang, W., Zhao, F., Ma, X., Perry, G. and Zhu, X. (2020) Mitochondria dysfunction in the pathogenesis of Alzheimer’s disease: recent advances. Mol Neurodegener 15, 30.

Wang, Y., Cella, M., Mallinson, K. et al. (2015) TREM2 lipid sensing sustains the microglial response in an Alzheimer’s disease model. Cell 160, 1061–1071.

Xie, L., Varathan, P., Nho, K., Saykin, A. J., Salama, P. and Yan, J. (2020) Identification of functionally connected multi-omic biomarkers for Alzheimer’s disease using modularity-constrained Lasso. PLoS One 15, e0234748.

Yang, D. S., Stavrides, P., Saito, M. et al. (2014) Defective macroautophagic turnover of brain lipids in the TgCRND8 Alzheimer mouse model: prevention by correcting lysosomal proteolytic deficits. Brain 137, 3300–3318.

Yu, H., Lin, X., Wang, D. et al. (2018) Mitochondrial Molecular Abnormalities Revealed by Proteomic Analysis of Hippocampal Organelles of Mice Triple Transgenic for Alzheimer Disease. Front Mol Neurosci 11, 74.

Zeisel, A., Muñoz-Manchado, A. B., Codeluppi, S. et al. (2015) Brain structure. Cell types in the mouse cortex and hippocampus revealed by single-cell RNA-seq. Science 347, 1138–1142.

Zerbino, D. R., Achuthan, P., Akanni, W. et al. (2018) Ensembl 2018. Nucleic Acids Res 46, D754–d761.

Zhang, X., Liu, W., Cao, Y. and Tan, W. (2020) Hippocampus Proteomics and Brain Lipidomics Reveal Network Dysfunction and Lipid Molecular Abnormalities in APP/PS1 Mouse Model of Alzheimer’s Disease. J Proteome Res 19, 3427–3437.

Zhou, Q., Liu, M., Xia, X. et al. (2017) A mouse tissue transcription factor atlas. Nat Commun 8, 15089.

