## Supplementary Table 1 for "Pathway-based integration of multi-omics data reveals lipidomics alterations validated in an Alzheimer’s Disease mouse model and risk loci carriers"

**Supplementary Table 1.** Transcription Factor enrichment analysis of all-mapped AD transcriptomics and proteomics datasets

| Transcription factor | Gene or protein target coverage | Fishers exact test p-value | FDR adj.  p-value | Odds Ratio |
| --- | --- | --- | --- | --- |
| Transcriptomics | **all** |  |  |  |
| CTCF | 83/1646 | 4.28e-07 | 2.36e-04 | 1.906 |
| MAFK | 76/1616 | 1.20e-50 | 0.0013 | 1.772 |
| MAZ | 78/1766 | 6.92e-05 | 0.0029 | 1.659 |
| CEBPB | 60/1285 | 9.48e-05 | 0.0035 | 1.758 |
| ETS1 | 71/1614 | 1.48e-04 | 0.0051 | 1.652 |
| MAX | 113/2892 | 3.24e-04 | 0.0089 | 1.46 |
| EGR1 | 73/1773 | 7.25e-04 | 0.016 | 1.542 |
| TAL1 | 77/1904 | 8.77e-04 | 0.019 | 1.513 |
| MYOD1 | 40/860 | 0.0011 | 0.023 | 1.751 |
| BHLHE40 | 70/1727 | 0.0013 | 0.025 | 1.517 |
| TCF3 | 69/1708 | 0.0015 | 0.026 | 1.511 |
| NFYA | 73/1834 | 0.0017 | 0.028 | 1.488 |
| TBP | 68/1687 | 0.0017 | 0.028 | 1.508 |
| USF2 | 71/1814 | 0.0027 | 0.032 | 1.462 |
| HNF4A | 89/2396 | 0.0040 | 0.042 | 1.385 |
| NR3C1 | 13/204 | 0.0043 | 0.043 | 2.443 |
| JUN | 53/1325 | 0.0053 | 0.048 | 1.496 |
| Proteomics | **all** |  |  |  |
| TAL1 | 35/690 | 5.41e-07 | 4.97e-05 | 2.759 |
| SREBF1 | 23/455 | 3.97e-05 | 8.36e-04 | 2.749 |
| SREBF2 | 15/366 | 0.0055 | 0.017 | 2.206 |
| BHLHE40 | 67/1702 | 2.12e-07 | 2.82e-05 | 2.116 |
| FOS | 45/1146 | 1.43e-05 | 4.66e-04 | 2.11 |
| SP4 | 53/1367 | 4.33e-06 | 1.99e-04 | 2.083 |
| REST | 70/1816 | 2.56e-07 | 2.82e-05 | 2.07 |
| MAX | 65/1728 | 1.31e-06 | 7.25e-05 | 2.018 |
| MXI1 | 66/1760 | 1.23e-06 | 7.25e-05 | 2.012 |
| CTCF | 68/1821 | 1.03e-06 | 7.25e-05 | 2.003 |
| NRF1 | 68/1827 | 1.14e-06 | 7.25e-05 | 1.996 |
| MYC | 64/1726 | 2.40e-06 | 1.21e-04 | 1.988 |
| MAFK | 43/1169 | 8.83e-05 | 0.0010 | 1.972 |
| ESRRA | 34/934 | 4.88e-04 | 0.0030 | 1.951 |
| MAZ | 62/1714 | 6.80e-06 | 2.89e-04 | 1.938 |
| CEBPB | 64/1836 | 1.49e-05 | 4.66e-04 | 1.865 |
| IRF3 | 58/1688 | 4.80e-05 | 8.36e-04 | 1.837 |
| ZBTB7A | 59/1735 | 5.52e-05 | 8.47e-04 | 1.818 |
| TBP | 62/1833 | 4.40e-05 | 8.36e-04 | 1.808 |
| CREB1 | 58/1731 | 9.07e-05 | 0.00104 | 1.79 |
| ETS1 | 54/1614 | 1.52e-04 | 0.0015 | 1.787 |
| EGR1 | 54/1627 | 1.83e-04 | 0.0015 | 1.772 |
| PAX5 | 27/817 | 0.0058 | 0.017 | 1.765 |
| E2F4 | 61/1847 | 9.22e-05 | 0.0010 | 1.763 |
| ATF3 | 42/1278 | 9.37e-04 | 0.0047 | 1.754 |
| USF1 | 81/2480 | 1.52e-05 | 4.66e-04 | 1.743 |
| RELA | 59/1815 | 1.71e-04 | 0.0015 | 1.735 |
| USF2 | 59/1820 | 1.82e-04 | 0.0015 | 1.73 |
| NFYB | 60/1852 | 1.66e-04 | 0.0015 | 1.729 |
| ZKSCAN1 | 60/1857 | 1.78e-04 | 0.0015 | 1.724 |
| NFYA | 59/1834 | 2.19e-04 | 0.0017 | 1.716 |
| BACH1 | 58/1814 | 2.81e-04 | 0.0020 | 1.705 |
| RFX5 | 58/1832 | 3.54e-04 | 0.0024 | 1.688 |
| YY1 | 84/2657 | 3.24e-05 | 0.00075 | 1.686 |
| TCF3 | 54/1708 | 5.43e-04 | 0.0032 | 1.686 |
| STAT1 | 58/1855 | 4.70e-04 | 0.0029 | 1.666 |
| TFAP2C | 56/1822 | 8.25e-04 | 0.0043 | 1.637 |
| SRF | 45/1467 | 0.0023 | 0.0088 | 1.634 |
| NR2C2 | 41/1343 | 0.0037 | 0.013 | 1.626 |
| NFE2 | 56/1854 | 0.0012 | 0.0057 | 1.608 |
| MYOD1 | 84/2784 | 1.35e-04 | 0.0013 | 1.606 |
| GATA3 | 30/997 | 0.013 | 0.030 | 1.602 |
| TCF12 | 30/1012 | 0.015 | 0.035 | 1.577 |
| PBX3 | 58/1958 | 0.0015 | 0.0068 | 1.576 |
| MYBL2 | 44/1487 | 0.0047 | 0.015 | 1.574 |
| STAT5A | 36/1230 | 0.011 | 0.026 | 1.557 |
| E2F6 | 71/2432 | 8.27e-04 | 0.0043 | 1.553 |
| SP1 | 47/1611 | 0.0047 | 0.015 | 1.552 |
| SP2 | 60/2063 | 0.0019 | 0.0077 | 1.547 |
| ARID3A | 53/1828 | 0.0034 | 0.012 | 1.542 |
| TEAD4 | 36/1245 | 0.013 | 0.029 | 1.537 |
| CTCFL | 62/2154 | 0.0021 | 0.0081 | 1.53 |
| JUND | 52/1821 | 0.0048 | 0.015 | 1.518 |
| EBF1 | 75/2676 | 0.0017 | 0.0073 | 1.489 |
| ELK1 | 51/1837 | 0.0084 | 0.022 | 1.474 |
| ELF1 | 52/1875 | 0.0080 | 0.022 | 1.473 |
| TFAP2A | 50/1810 | 0.0096 | 0.024 | 1.467 |
| POU2F2 | 43/1563 | 0.0156 | 0.035 | 1.461 |
| ATF1 | 50/1833 | 0.012 | 0.028 | 1.448 |
| GABPA | 62/2285 | 0.0068 | 0.019 | 1.44 |
| E2F1 | 50/1854 | 0.014 | 0.032 | 1.431 |
| TCF7L2 | 69/2574 | 0.0062 | 0.018 | 1.422 |
| HMGN3 | 49/1830 | 0.0167 | 0.036 | 1.421 |
| CUX1 | 48/1827 | 0.023 | 0.047 | 1.393 |
| IRF1 | 105/4204 | 0.0083 | 0.022 | 1.323 |
| STAT3 | 85/3450 | 0.019 | 0.040 | 1.304 |

FDR adj. refers to Benjamini-Hochberg (B-H) False Discovery Rate correction for multiple testing.
