## Supplementary Table 2 for "Pathway-based integration of multi-omics data reveals lipidomics alterations validated in an Alzheimer’s Disease mouse model and risk loci carriers"

**Supplementary Table 2.** Biological Process (BP), Molecular Function (MF) and Cellular Compartment (CC) enrichment analysis of all-mapped AD transcriptomics, proteomics and GWAS-orthologs datasets

| GO ID | Term | Term  Coverage  (mapped/all) | Fold  Enrichment | P value | FDR adj.  p value |
| --- | --- | --- | --- | --- | --- |
|  | **Transcriptomics (BP)** |  |  |  |  |
| GO:0016310 | Phosphorylation | 105/612 | 5.36 | 1.32e-46 | 2.34e-43 |
| GO:0006468 | Protein phosphorylation | 90/576 | 4.88 | 2.19e-36 | 3.88e-33 |
| GO:0055114 | Oxidation-reduction process | 95/676 | 4.39 | 1.23e-34 | 2.18e-31 |
| GO:0008152 | Metabolic process | 74/463 | 4.99 | 2.01e-30 | 3.57e-27 |
| GO:0006629 | Lipid metabolic process | 72/459 | 4.90 | 4.54e-29 | 8.05e-26 |
| GO:0046777 | Protein autophosphorylation | 46/183 | 7.85 | 2.15e-27 | 3.81e-24 |
| GO:0035556 | Intracellular signal transduction | 51/400 | 3.98 | 9.65e-17 | 2.00e-13 |
| GO:0006631 | Fatty acid metabolic process | 30/156 | 6.00 | 1.36e-14 | 2.42e-11 |
| GO:0018105 | Peptidyl-serine phosphorylation | 26/133 | 6.10 | 7.13e-13 | 1.27e-09 |
| GO:0018108 | Peptidyl-tyrosine phosphorylation | 18/65 | 8.65 | 1.35e-11 | 2.40e-08 |
| GO:0005975 | Carbohydrate metabolic process | 28/206 | 4.24 | 5.09e-10 | 9.03e-07 |
| GO:0006198 | cAMP catabolic process | 9/16 | 17.56 | 1.07e-8 | 1.89e-05 |
| GO:0006811 | Ion transport | 47/568 | 2.51 | 1.77e-08 | 3.13e-05 |
| GO:0038083 | Peptidyl-tyrosine autophosphorylation | 12/41 | 9.14 | 4.28e-08 | 7.60e-05 |
| GO:0006470 | Protein dephosphorylation | 20/138 | 4.53 | 8.56e-08 | 1.52e-04 |
| GO:0016126 | Sterol biosynthetic process | 10/27 | 11.566 | 9.17e-08 | 1.63e-04 |
| GO:0006694 | Steroid biosynthetic process | 13/63 | 6.55 | 5.10e-07 | 9,05e-4 |
| GO:0007169 | Transmembrane receptor protein tyrosine kinase signaling pathway | 16/100 | 4.99 | 6.40e-07 | 0.0011 |
| GO:0016042 | Lipid catabolic process | 16/109 | 4.58 | 1.97e-06 | 0.0035 |
| GO:0016311 | Dephosphorylation | 16/109 | 4.58 | 1.97e-06 | 0.0035 |
| GO:0055085 | Transmembrane transport | 31/364 | 2.66 | 2.28e-06 | 0.0040 |
| GO:0006749 | Glutathione metabolic process | 11/49 | 7.01 | 2.75e-06 | 0.0048 |
| GO:0006695 | Cholesterol biosynthetic process | 9/32 | 8.78 | 5.54e-06 | 0.009 |
| GO:0010033 | Response to organic substance | 12/65 | 5.76 | 6.05e-06 | 0.010 |
| GO:0034765 | Regulation of ion transmembrane transport | 17/135 | 3.93 | 6.61e-06 | 0.011 |
| GO:0006635 | Fatty acid beta-oxidation | 10/44 | 7.09 | 8.54e-06 | 0.015 |
| GO:0042493 | Response to drug | 28/339 | 2.57 | 1.36e-05 | 0.024 |
| GO:0008202 | Steroid metabolic process | 13/84 | 4.83 | 1.41e-05 | 0.025 |
| GO:0009636 | Response to toxic substance | 13/86 | 4.72 | 1.80e-05 | 0.032 |
| GO:0006486 | Protein glycosylation | 15/116 | 4.04 | 2.01e-05 | 0.035 |
|  | **Proteomics (BP)** |  |  |  |  |
| GO:0055114 | Oxidation-reduction process | 74/676 | 5.08 | 4.82e-31 | 8.27e-28 |
| GO:0008152 | Metabolic process | 59/463 | 5.90 | 5.11e-28 | 8.77e-25 |
| GO:0016310 | Phosphorylation | 57/612 | 4.31 | 2.32e-20 | 3.98e-17 |
| GO:0006629 | Lipid metabolic process | 44/459 | 4.44 | 4.54e-16 | 7.66e-13 |
| GO:0006468 | Protein phosphorylation | 46/576 | 3.70 | 6.68e-14 | 1.14e-10 |
| GO:0006099 | Tricarboxylic acid cycle | 12/29 | 19.18 | 9.70e-12 | 1.66e-08 |
| GO:0008652 | Cellular amino-acid biosynthetic process | 9/25 | 16.69 | 3.35e-08 | 5.75e-05 |
| GO:0006631 | Fatty acid metabolic process | 18/156 | 5.35 | 4.57e-08 | 7.85e-05 |
| GO:0005975 | Carbohydrate metabolic process | 20/206 | 4.50 | 1.10e-07 | 1.89e-04 |
| GO:0045454 | Cell redox homeostasis | 12/65 | 8.56 | 1.25e-07 | 2.15e-04 |
| GO:0006470 | Protein dephosphorylation | 16/138 | 5.37 | 2.94e-07 | 5.05e-04 |
| GO:0006749 | Glutathione metabolic process | 10/49 | 9.46 | 8.64e-07 | 0.0015 |
| GO:0035556 | Intracellular signal transduction | 25/400 | 2.89 | 6.45e-06 | 0.011 |
| GO:0046777 | Protein autophosphorylation | 16/183 | 4.05 | 1.03e-05 | 0.017 |
| GO:0009058 | Biosynthetic process | 8/37 | 10.02 | 1.19e-05 | 0.021 |
| GO:0006006 | Glucose metabolic process | 10/68 | 6.81 | 1.45e-05 | 0.025 |
| GO:0016311 | Dephosphorylation | 12/109 | 5.10 | 2.31e-05 | 0.039 |
|  | **GWAS-orthologs (BP)** |  |  |  |  |
| GO:0016310 | Phosphorylation | 50/612 | 5.75 | 2.16e-23 | 3.56e-20 |
| GO:0006468 | Protein phosphorylation | 44/576 | 5.37 | 2.08e-19 | 3.43e-06 |
| GO:0046777 | Protein autophosphorylation | 19/183 | 7.30 | 1.25e-10 | 2.07e-07 |
| GO:0006811 | Ion transport | 27/584 | 3.25 | 2.63e-07 | 4.35e-04 |
| GO:0006470 | Protein dephosphorylation | 13/138 | 6.63 | 6.30e-07 | 0.0010 |
| GO:0008152 | Metabolic process | 23/463 | 3.50 | 7.86e-07 | 0.0013 |
| GO:0055085 | Transmembrane transport | 20/364 | 3.86 | 1.15e-06 | 0.0019 |
| GO:0034765 | Ion transmembrane transport regulation | 12/135 | 6.25 | 3.52e-06 | 0.0058 |
| GO:0055114 | Oxidation-reduction process | 27/676 | 2.81 | 4.05e-06 | 0.0067 |
| GO:0006629 | Lipid metabolic process | 21/459 | 3.21 | 9.22e-06 | 0.015 |
| GO:0018105 | Peptidyl-serine phosphorylation | 11/133 | 5.82 | 1.99e-05 | 0.033 |
|  | **Transcriptomics (MF)** |  |  |  |  |
| GO:0016740 | Transferase activity | 191/1472 | 3.84 | 4.04e-64 | 6.42e-61 |
| GO:0016301 | Kinase activity | 109/674 | 4.79 | 9.91e-44 | 1.58e-40 |
| GO:0004672 | Protein kinase activity | 83/531 | 4.63 | 7.05e-32 | 1.12e-28 |
| GO:0016491 | Oxidoreductase activity | 88/604 | 4.31 | 1.63e-31 | 2.60e-28 |
| GO:0005524 | ATP binding | 142/1507 | 2.79 | 2.60e-30 | 4.14e-27 |
| GO:0000166 | Nucleotide binding | 157/1936 | 2.40 | 1.66e-26 | 2.63e-23 |
| GO:0016787 | Hydrolase activity | 136/1533 | 2.62 | 2.43e-26 | 3.88e-23 |
| GO:0003824 | Catalytic activity | 69/479 | 4.27 | 2.50e-24 | 3.98e-21 |
| GO:0004674 | Protein serine/threonine kinase activity | 63/428 | 4.36 | 1.03e-22 | 1.64e-19 |
| GO:0004713 | Protein tyrosine kinase activity | 26/121 | 6.36 | 2.40e-13 | 3.81e-10 |
| GO:0008081 | Phosphoric diester hydrolase activity | 15/52 | 8.54 | 1.14e-09 | 1.81e-06 |
| GO:0051287 | NAD binding | 15/55 | 8.08 | 2.55e-09 | 4.05e-06 |
| GO:0004721 | Phosphoprotein phosphatase activity | 22/139 | 4.67 | 8.29e-09 | 1.32e-05 |
| GO:0016746 | Transferase activity, transferring acyl groups | 24/167 | 4.26 | 1.00e-08 | 1.59e-05 |
| GO:0004715 | Non-membrane spanning prot. tyrosine kinase act. | 13/45 | 8.55 | 2.00e-08 | 3.18e-05 |
| GO:0016829 | Lyase activity | 21/139 | 4.48 | 4.37e-08 | 6.95e-04 |
| GO:0004029 | Aldehyde dehydrogenase (NAD) activity | 9/19 | 14.03 | 8.64e-08 | 1.38e-04 |
| GO:0016757 | Transferase activity, transf. glycosyl groups | 25/208 | 3.56 | 1.54e-07 | 2.45e-04 |
| GO:0016791 | Phosphatase activity | 18/118 | 4.52 | 4.40e-07 | 7.00e-04 |
| GO:0046872 | Metal ion binding | 162/3355 | 1.43 | 6.58e-07 | 0.0010 |
| GO:0004114 | 3',5'-cyclic-nucleotide phosphodiesterase activity | 9/24 | 11.11 | 7.24e-07 | 0.0012 |
| GO:0004725 | Protein tyrosine phosphatase activity | 16/97 | 4.88 | 8.29e-07 | 0.0013 |
| GO:0005536 | Glucose binding | 7/12 | 17.38 | 1.11e-06 | 0.0018 |
| GO:0008234 | Cysteine-type peptidase activity | 19/141 | 3.99 | 1.26e-06 | 0.0020 |
| GO:0004714 | Transmembrane recept. protein tyrosine kinase act. | 12/54 | 6.58 | 1.49e-06 | 0.0023 |
| GO:0004115 | 3',5'-cyclic-AMP phosphodiesterase activity | 7/13 | 15.94 | 2.00e-06 | 0.0032 |
| GO:0004602 | Glutathione peroxidase activity | 8/20 | 11.85 | 2.52e-06 | 0.0040 |
| GO:0015293 | Symporter activity | 16/112 | 4.23 | 5.28e-06 | 0.0084 |
| GO:0016853 | Isomerase activity | 16/115 | 4.12 | 7.34e-06 | 0.011 |
| GO:0016620 | Oxidoreductase activity, acting on the aldehyde or oxo group of donors | 9/32 | 8.33 | 8.16e-06 | 0.013 |
| GO:0004364 | Glutathione transferase activity | 9/33 | 8.08 | 1.05e-05 | 0.016 |
| GO:0005244 | Voltage-gated ion channel activity | 17/134 | 3.76 | 1.16e-05 | 0.018 |
| GO:0000287 | Magnesium ion binding | 21/202 | 3.08 | 1.70e-05 | 0.027 |
| GO:0030552 | cAMP binding | 8/26 | 9.11 | 1.79e-05 | 0.028 |
|  | **Proteomics (MF)** |  |  |  |  |
| GO:0016740 | Transferase activity | 105/1472 | 3.25 | 4.27e-28 | 6.59e-25 |
| GO:0003824 | Catalytic activity | 60/479 | 5.71 | 9.72e-28 | 1.50e-24 |
| GO:0016787 | Hydrolase activity | 99/1533 | 2.94 | 4.30e-23 | 6.64e-20 |
| GO:0005524 | ATP binding | 98/1507 | 2.96 | 4.63e-23 | 7.26e-20 |
| GO:0000166 | Nucleotide binding | 111/1936 | 2.61 | 5.49e-22 | 8.49e-19 |
| GO:0016491 | Oxidoreductase activity | 59/604 | 4.45 | 9.33e-22 | 1.44e-18 |
| GO:0016301 | Kinase activity | 61/674 | 4.12 | 7.73e-21 | 1.19e-17 |
| GO:0004672 | Protein kinase activity | 43/531 | 3.67 | 5.61e-13 | 8.66e-10 |
| GO:0004674 | Protein serine/threonine kinase activity | 36/428 | 3.83 | 2.13e-11 | 3.29e-08 |
| GO:0051287 | NAD binding | 12/55 | 9.94 | 2.42e-08 | 3.74e-05 |
| GO:0016853 | Isomerase activity | 16/115 | 6.33 | 3.18e-08 | 4.91e-05 |
| GO:0004702 | Receptor sig. protein serine/threonine kinase act. | 12/57 | 9.58 | 3.59e-08 | 5.55e-05 |
| GO:0004721 | Phosphoprotein phosphatase activity | 17/139 | 5.57 | 6.66e-08 | 1.03e-04 |
| GO:0016829 | Lyase activity | 17/139 | 5.57 | 6.66e-08 | 1.03e-04 |
| GO:0008081 | Phosphoric diester hydrolase activity | 11/52 | 9.63 | 1.57e-07 | 2.43e-04 |
| GO:0000287 | Magnesium ion binding | 18/202 | 4.06 | 2.34e-06 | 0.0036 |
| GO:0042803 | Protein homodimerization activity | 40/798 | 2.28 | 2.42e-06 | 0.0037 |
| GO:0030170 | Pyridoxal phosphate binding | 10/55 | 8.28 | 2.75e-06 | 0.0042 |
| GO:0019901 | Protein kinase binding | 27/434 | 2.83 | 3.81e-06 | 0.0059 |
| GO:0004725 | Protein tyrosine phosphatase activity | 12/97 | 5.63 | 8.86e-06 | 0.014 |
| GO:0008137 | NADH dehydrogenase (ubiquinone) activity | 8/35 | 10.41 | 9.05e-06 | 0.014 |
| GO:0016791 | Phosphatase activity | 13/118 | 5.01 | 1.08e-05 | 0.017 |
| GO:0019899 | Enzyme binding | 24/384 | 2.84 | 1.39e-05 | 0.021 |
| GO:0016874 | Ligase activity | 23/362 | 2.89 | 1.71e-05 | 0.026 |
| GO:0009055 | Electron carrier activity | 9/53 | 7.74 | 1.84e-05 | 0.028 |
|  | **GWAS-orthologs (MF)** |  |  |  |  |
| GO:0016740 | Transferase activity | 93/1472 | 4.30 | 1.94e-35 | 2.87e-32 |
| GO:0016301 | Kinase activity | 52/674 | 5.25 | 1.22e-22 | 1.80e-19 |
| GO:0005524 | ATP binding | 74/1507 | 3.34 | 7.27e-21 | 1.07e-17 |
| GO:0004672 | Protein kinase activity | 42/531 | 5.40 | 1.40e-18 | 2.07e-15 |
| GO:0000166 | Nucleotide binding | 77/1936 | 2.71 | 2.73e-16 | 3.33e-13 |
| GO:0004674 | Protein serine/threonine kinase activity | 34/428 | 5.41 | 5.10e-15 | 7.54e-12 |
| GO:0016829 | Lyase activity | 16/139 | 7.84 | 1.91e-09 | 2.83e-06 |
| GO:0003824 | Catalytic activity | 28/479 | 3.98 | 2.16e-09 | 3.18e-06 |
| GO:0046872 | Metal ion binding | 88/3355 | 1.78 | 1.16e-08 | 1.71e-05 |
| GO:0004721 | Phosphoprotein phosphatase activity | 14/139 | 6.86 | 1.30e-07 | 1.92e-04 |
| GO:0016791 | Phosphatase activity | 12/118 | 6.93 | 1.26e-06 | 0..0019 |
| GO:0000287 | Magnesium ion binding | 15/202 | 5.06 | 1.69e-06 | 0.0025 |
| GO:0005244 | Voltage-gated ion channel activity | 12/134 | 6.10 | 4.43e-06 | 0.0065 |
| GO:0016853 | Isomerase activity | 11/115 | 6.51 | 7.24e-06 | 0.011 |
| GO:0016491 | Oxidoreductase activity | 25/604 | 2.82 | 9.43e-06 | 0.013 |
| GO:0004812 | Aminoacyl-tRNA ligase activity | 7/39 | 12.23 | 2.00e-05 | 0.029 |
| GO:0016787 | Hydrolase activity | 44/1533 | 1.95 | 2.41e-05 | 0.035 |
|  | **Transcriptomics (CC)** |  |  |  |  |
| GO:0005829 | Cytosol | 136/1784 | 2.63 | 2.11e-26 | 2.92e-23 |
| GO:0070062 | Extracellular exosome | 167/2674 | 2.15 | 3.07e-23 | 4.26e-20 |
| GO:0016020 | Membrane | 313/6998 | 1.54 | 6.74e-22 | 9.35e-19 |
| GO:0005739 | Mitochondrion | 121/1721 | 2.43 | 2.22e-20 | 3.09e-17 |
| GO:0005737 | Cytoplasm | 266/6631 | 1.39 | 6.73e-11 | 9.34e-08 |
| GO:0005783 | Endoplasmic reticulum | 81/1323 | 2.11 | 2.23e-10 | 3.10e-07 |
| GO:0005764 | Lysosome | 33/331 | 3.44 | 2.60e-09 | 3.61e-06 |
| GO:0005759 | Mitochondrial matrix | 22/188 | 4.04 | 1.29e-07 | 1.79e-04 |
| GO:0045121 | Membrane raft | 26/262 | 3.43 | 1.87e-07 | 2.60e-04 |
| GO:0005743 | Mitochondrial inner membrane | 32/387 | 2.86 | 3.43e-07 | 4.77e-04 |
| GO:0005789 | Endoplasmic reticulum membrane | 45/710 | 2.19 | 1.80e-06 | 0.0025 |
| GO:0016323 | Basolateral plasma membrane | 21/203 | 3.57 | 1.92e-06 | 0.0026 |
| GO:0005794 | Golgi apparatus | 64/1190 | 1.86 | 2.39e-06 | 0.0033 |
| GO:0043025 | Neuronal cell body | 37/534 | 2.39 | 2.45e-06 | 0.0034 |
| GO:0031234 | Extrinsic – plasma membrane cytoplasmic side | 12/68 | 6.09 | 3.66e-06 | 0.0051 |
| GO:0043005 | Neuron projection | 31/420 | 2.55 | 5.52e-06 | 0.0076 |
| GO:0043231 | Intracellular membrane-bounded organelle | 45/751 | 2.07 | 7.56e-06 | 0.010 |
| GO:0048471 | Perinuclear region of cytoplasm | 41/692 | 2.04 | 2.70e-05 | 0.037 |
| GO:0005887 | Plasma membrane | 58/1126 | 1.77 | 2.71e-05 | 0.037 |
| GO:0043204 | Perikaryon | 16/250 | 3.68 | 3.04e-05 | 0.042 |
| GO:0016324 | Apical plasma membrane | 25/328 | 2.63 | 3.23e-05 | 0.045 |
|  | **Proteomics (CC)** |  |  |  |  |
| GO:0005739 | Mitochondrion | 147/1721 | 4.36 | 4.68e-57 | 6.33e-54 |
| GO:0070062 | Extracellular exosome | 146/2674 | 2.78 | 3.45e-33 | 4.66e-30 |
| GO:0005743 | Mitochondrial inner membrane | 43/387 | 5.67 | 1.24e-19 | 1.67e-16 |
| GO:0005829 | Cytosol | 93/1784 | 2.66 | 1.30e-18 | 1.76e-15 |
| GO:0005737 | Cytoplasm | 206/6631 | 1.58 | 1.06e-15 | 1.50e-12 |
| GO:0005747 | Mitochondrial res. chain complex I | 16/47 | 17.38 | 7.49e-15 | 1.00e-11 |
| GO:0070469 | Respiratory chain | 17/58 | 14.96 | 1.24e-14 | 1.68e-11 |
| GO:0043209 | Myelin sheath | 24/192 | 6.38 | 3.99e-12 | 5.39e-09 |
| GO:0005759 | Mitochondrial matrix | 23/188 | 6.24 | 1.89e-11 | 2.55e-08 |
| GO:0016020 | Membrane | 195/6998 | 1.43 | 1.06e-09 | 1.43e-06 |
| GO:0005777 | Peroxisome | 15/131 | 5.85 | 2.87e-07 | 3.88e-04 |
| GO:0022624 | Proteasome accessory complex | 7/17 | 21.02 | 5.51e-07 | 7.44e-04 |
| GO:0043231 | Intracellular membrane-bounded organelle | 37/751 | 2.51 | 7.21e-07 | 9.74e-04 |
| GO:0043005 | Neuron projection | 25/420 | 3.04 | 3.00e-06 | 0.0041 |
| GO:0008540 | Proteasome regulatory particle I | 6/13 | 23.57 | 3.13e-06 | 0.0040 |
| GO:0000502 | Proteasome complex | 10/66 | 7.73 | 5.22e-06 | 0.0070 |
| GO:0005891 | Voltage-gated calcium channel complex | 7/24 | 14.89 | 5.33e-06 | 0.0072 |
| GO:0005778 | Peroxisomal membrane | 9/23 | 8.67 | 8.09e-06 | 0.011 |
| GO:0005838 | Proteasome regulatory particle II | 5/10 | 25.53 | 2.74e-05 | 0.037 |
| GO:0031597 | Cytosolic proteasome complex | 5/10 | 25.53 | 2.74e-05 | 0.037 |
| GO:0005749 | Mitochondrial respiratory chain complex I | 4/4 | 51.07 | 2.91e-05 | 0.039 |
|  | **GWAS-orthologs (CC)** |  |  |  |  |
| GO:0016020 | Membrane | 136/6998 | 1.51 | 3.42e-09 | 4.45e-06 |

FDR adj. refers to Benjamini-Hochberg (B-H) False Discovery Rate correction for multiple testing.
