## Supplementary Table 3 for "Pathway-based integration of multi-omics data reveals lipidomics alterations validated in an Alzheimer’s Disease mouse model and risk loci carriers"

**Supplementary Table 3.** Transcription Factor enrichment analysis of AD-metabolic transcriptomics and proteomics datasets

| Transcription factor | Gene or protein target coverage | Fishers exact test p-value | FDR adj.  p-value | Odds Ratio |
| --- | --- | --- | --- | --- |
| AD-metabolic | **Transcriptomics** |  |  |  |
| ESRRA | 8/108 | 1.16e-05 | 0.00321 | 8.341 |
| SREBF1 | 17/455 | 4.15e-06 | 0.00229 | 4.047 |
| HNF4G | 16/556 | 1.62e-04 | 0.0297 | 3.089 |
| TAL1 | 19/820 | 6.34e-04 | 0.05 | 2.473 |
| CTCF | 38/2137 | 6.05e-3 | 0.05 | 1.888 |
| MAX | 49/2892 | 3.90e-4 | 0.05 | 1.797 |
| AD-metabolic | **Proteomics** |  |  |  |
| ESRRA | 7/108 | 3.31e-05 | 0.0013 | 8.594 |
| NFE2 | 10/258 | 6,72e-05 | 0.0019 | 5 |
| SREBF1 | 14/455 | 3.65e-05 | 0.0013 | 3.937 |
| SP4 | 32/1367 | 5.77e-07 | 2.95e-04 | 2.972 |
| TAL1 | 16/690 | 2.81e-04 | 0.0039 | 2.944 |
| CEBPB | 24/1083 | 2.57e-05 | 0.0013 | 2.81 |
| SREBF2 | 8/366 | 0.011 | 0.037 | 2.771 |
| MAX | 36/1719 | 1.65e-06 | 3.03e-04 | 2.652 |
| JUND | 12/598 | 0.0045 | 0.020 | 2.539 |
| CREB1 | 20/1013 | 4.45e-04 | 0.0047 | 2.497 |
| ATF3 | 25/1278 | 1.22e-04 | 0.0029 | 2.474 |
| NRF1 | 35/1827 | 1.33e-05 | 0.0011 | 2.422 |
| USF1 | 47/2480 | 1.07e-06 | 2.95e-04 | 2.395 |
| USF2 | 34/1820 | 2.72e-05 | 0.0013 | 2.361 |
| MAZ | 32/1714 | 4.40e-05 | 0.0014 | 2.359 |
| MYC | 34/1830 | 3.02e-05 | 0.0013 | 2.347 |
| BHLHE40 | 31/1702 | 8.50e-05 | 0.0022 | 2.3 |
| NR2C2 | 24/1343 | 5.57e-04 | 0.0053 | 2.256 |
| MXI1 | 31/1760 | 1.49e-04 | 0.0032 | 2.223 |
| CTCF | 32/1825 | 1.30e-04 | 0.003 | 2.213 |
| EGR1 | 31/1773 | 1.68e-04 | 0.0032 | 2.207 |
| ZEB1 | 14/803 | 0.0077 | 0.029 | 2.2 |
| TBP | 32/1843 | 1.54e-04 | 0.0032 | 2.191 |
| IRF3 | 32/1865 | 1.87e-04 | 0.0033 | 2.165 |
| ETS1 | 28/1661 | 5.38e-04 | 0.0052 | 2.126 |
| E2F4 | 31/1847 | 3.24e-04 | 0.0040 | 2.117 |
| NFYB | 31/1852 | 3.38e-04 | 0.0041 | 2.111 |
| MAFK | 27/1616 | 7.40e-04 | 0.0065 | 2.107 |
| ELF1 | 31/1875 | 4.10e-04 | 0.0046 | 2.085 |
| TFAP2C | 30/1822 | 5.24e-04 | 0.0052 | 2.076 |
| PAX5 | 34/2072 | 2.73e-04 | 0.0039 | 2.069 |
| CTCFL | 35/2154 | 2.72e-04 | 0.0039 | 2.048 |
| NFYA | 30/1873 | 7.91e-04 | 0.0067 | 2.018 |
| RFX5 | 29/1851 | 0.0013 | 0.0084 | 1.974 |
| SP1 | 25/1611 | 0.0028 | 0.014 | 1.955 |
| SP2 | 32/2063 | 9.27e-04 | 0.0072 | 1.954 |
| TFAP2A | 28/1810 | 0.0019 | 0.011 | 1.948 |
| RELA | 28/1815 | 0.0019 | 0.011 | 1.943 |
| REST | 28/1816 | 0.0019 | 0.011 | 1.942 |
| E2F6 | 37/2432 | 6.12e-04 | 0.0056 | 1.916 |
| MYOD1 | 42/2784 | 3.64e-04 | 0.0042 | 1.899 |
| NFIC | 16/1061 | 0.0164 | 0.046 | 1.899 |
| GABPA | 28/1864 | 0.00263 | 0.014 | 1.891 |
| BACH1 | 27/1814 | 0.0034 | 0.017 | 1.874 |
| FOS | 54/3683 | 1.58e-04 | 0.0032 | 1.845 |
| ZBTB7A | 25/1735 | 0.0066 | 0.026 | 1.813 |
| PBX3 | 28/1958 | 0.0049 | 0.021 | 1.799 |
| HNF4A | 34/2396 | 0.0026 | 0.014 | 1.785 |
| STAT1 | 26/1855 | 0.0080 | 0.029 | 1.763 |
| ZKSCAN1 | 26/1857 | 0.0081 | 0.029 | 1.761 |
| ATF1 | 25/1833 | 0.0121 | 0.038 | 1.715 |
| ELK1 | 25/1837 | 0.012 | 0.038 | 1.711 |
| TCF7L2 | 35/2574 | 0.0044 | 0.020 | 1.709 |
| YY1 | 36/2657 | 0.0042 | 0.020 | 1.703 |
| STAT3 | 42/3450 | 0.012 | 0.037 | 1.528 |
| IRF1 | 50/4204 | 0.011 | 0.036 | 1.493 |

FDR adjusted refers to Benjamini-Hochberg (B-H) False Discovery Rate correction for multiple testing.
