## Supplementary Table 4 for "Pathway-based integration of multi-omics data reveals lipidomics alterations validated in an Alzheimer’s Disease mouse model and risk loci carriers"

**Supplementary Table 4.** Metabolic pathway enrichment analysis of AD-metabolic transcriptomics, proteomics and GWAS-orthologs datasets

| **Pathway** | **Transcr/**  **protein/**  **gene cover** | **Transcripts/proteins/genes mapped** | **P value**  **(raw)** | **BH-adj**  **P value** |
| --- | --- | --- | --- | --- |
| **Transcriptomics** |  |  |  |  |
| Super-pathway of cholesterol biosynthesis | 9/25 | *Acat2, Dhcr24, Fdft1, Fdps, Hmgcs1, Hsd17b7, Nsdhl, Pmvk, Sqle* | 8.72e-6 | 1.21e-3 |
| Fatty acid α-oxidation | 5/10 | *Aldh1a7, Aldh3a1, Aldh3b1 Aldh7a1, Aldh9a1,* | 1.60e-4 | 8.58e-3 |
| Sucrose degradation | 4/6 | *Gpi, Hk1, Hk2, Hk3* | 1.85e-4 | 8.58e-3 |
| Fatty acid β-oxidation I | 8/31 | *Acaa2, Acadsb, Acadvl, Acsl5, Hadh, Hadhb, Peci, Slc27a2* | 3.97e-4 | 1.05e-2 |
| Putrescine degradation III | 4/7 | *Aldh1a7, Aldh7a1, Aldh9a1, Maob* | 4.12e-4 | 1.05e-2 |
| Cholesterol biosynthesis I | 5/12 | *Dhcr24, Fdft1, Hsd17b7, Nsdhl, Sqle* | 4.54e-4 | 1.05e-2 |
| Fatty acid β-oxidation IV | 4/8 | *Decr1, Decr2, Hadhb, Peci* | 7.84e-4 | 1.56e-2 |
| Tryptophan degradation III | 5/15 | *Acat2, Haao, Hadh, Ogdh, Tdo2* | 1.48e-3 | 2.57e-2 |
| Biosynthesis of prostaglandins | 4/11 | *Hpgd, Ptgds2, Ptgs1, Tbxas* | 3.19e-3 | 4.44e-2 |
| Cholesterol biosynthesis III | 4/11 | *Fdft1, Hsd17b7, Nsdhl, Sqle* | 3.19e-3 | 4.44e-2 |
| Epoxysqualene biosynthesis | 2/2 | *Fdft1, Sqle* | 3.73e-3 | 3.74e-2 |
| Fatty acid biosynthesis initiation III | 2/2 | *Fasn, Oxsm* | 3.73e-3 | 3.74e-2 |
| Fatty acid biosynthesis initiation II | 2/2 | *Fasn, Oxsm* | 3.73e-3 | 3.74e-2 |
| Phospholipases | 7/34 | *Pla2g4a, Pla2g4e, Pla2g5, Plce1, Plcg2, Plch1, Pld4* | 3.73e-3 | 3.74e-2 |
| **Proteomics** |  |  |  |  |
| Aerobic respiration – electron donor II | 21/72 | *Ndufa11, Ndufa12, Ndufa3, Ndufa4, Ndufa5, Ndufa7, Ndufa9, Ndufab1, Ndufb11, Ndufb6, Ndufb7, Ndufs5, Ndufs6, Ndufs7, Ndufs8, Ndufv2, Sdha, Sdhb, Sdhc, Sdhd, Uqcrfs1* | 9.26e-12 | 1.31e-9 |
| NADH to cytochrome bo oxidase electron transfer | 16/51 | *Ndufa11, Ndufa 12, Ndufa3, Ndufa4, Ndufa5, Ndufa7, Ndufa9, Ndufab1, Ndufb11, Ndufb6, Ndufb7, Ndufs5, Ndufs6, Ndufs7, Ndufs8, Ndufv2* | 9.78e-10 | 4.59e-8 |
| NADH to cytochrome bd oxidase electron transfer | 16/51 | *Same enzymes* as NADH to cytochrome bo oxidase electron transfer | 9.78e-10 | 4.59e-8 |
| TCA cycle | 8/13 | *Aco1, Idh2, Sdha, Sdhb, Sdhc, Sdhd, Suclg1, Suclg2* | 3.14e-8 | 1.11e-6 |
| TCA cycle variation III | 8/14 | *Aco1, Idh3a, Sdha, Sdhb, Sdhc, Sdhd, Suclg1, Suclg2* | 7.02e-8 | 1.98e-6 |
| Aerobic respiration - electron donors reaction list | 5/7 | *Gpd1, Sdha, Sdhb, Sdhc, Sdhd* | 5.39e-6 | 1.27e-4 |
| Heme degradation | 3/4 | *Blvra, Blvrb, Hmox2* | 4.57e-4 | 9.21e-3 |
| Gluconeogenesis I | 5/21 | *Gapdh, Me1, Me3, Pgam1, Pgam2* | 2.96 e-3 | 2.61e-2 |
| Phosphatidyl-  glycerol biosynthesis I | 4/13 | *Agpat1, Agpat4, Csd2, Lpcat4* | 2.90 e-3 | 2.61e-2 |
| CDP-diglycerol biosynthesis I | 4/13 | *Same enzymes* as phosphatidyl-  glycerol biosynthesis I | 2.90e-3 | 2.61e-2 |
| Aspartate biosynthesis | 2/2 | *Got2, Pcx* | 2.43e-3 | 2.63e-2 |
| Cysteine biosynthesis/  Homocysteine degradation | 2/2 | *Cbs, Cth* | 2.43e-3 | 2.63e-2 |
| Cysteine biosynthesis II | 2/2 | *Same enzymes as* Cysteine biosynthesis/  Homocysteine degradation | 2.43e-3 | 2.63e-2 |
| Super-pathway of acetyl-CoA biosynthesis | 3/6 | *Acly, Dlat, Pdha1* | 2.12e-3 | 2.99e-2 |
| Glycolysis V | 4/12 | *Adpgk, Pgam1, Pgam2, Tpi1* | 2.09e-3 | 2.99e-2 |
| CDP-diglycerol biosynthesis II | 4/12 | *Same enzymes* as phosphatidyl-  glycerol biosynthesis I | 2.09e-3 | 2.99e-2 |
| Phosphatidyl-  glycerol biosynthesis II | 4/4 | *Same enzymes* as phosphatidyl-  glycerol biosynthesis I | 3.90e-3 | 3.24e-2 |
| **GWAS-orthologs** |  |  |  |  |
| Thyroid hormone metabolism I | 2/3 | *Dio1, Dio3* | 8.91e-4 | 3.44e-2 |
| Aerobic respiration – electron donor II | 6/72 | *Ndufa12, Ndufa2, Sdhb, Ndufc1, Ndufs3, Ndufs7* | 1.40e-3 | 3.44e-2 |
| NADH to cytochrome bo oxidase electron transfer | 5/51 | *Ndufa12, Ndufa2, Ndufc1, Ndufs3, Ndufs7* | 1.74e-3 | 3.44e-2 |
| NADH to cytochrome bd oxidase electron transfer | 5/51 | *Same enzymes* as NADH to cytochrome bo oxidase electron transfer | 1.74e-3 | 3.44e-2 |

B-H adj. refers to Benjamini-Hochberg (B-H) FDR-adjusted p-value. Underlined genes/transcripts/proteins refer to omics elements that were mapped onto more than one metabolic pathway.
