## Supplementary Table 5 for "Pathway-based integration of multi-omics data reveals lipidomics alterations validated in an Alzheimer’s Disease mouse model and risk loci carriers"

**Supplementary Table 5.** Unconditional and conditional EWCE analysis of AD-metabolic multi-omics datasets

| Cell type | Fold change | SD  from the mean | P value | B-H adj.  p value | Condition |
| --- | --- | --- | --- | --- | --- |
| Transcriptomics | **dataset** |  |  |  |  |
| Astrocytes (ependymal) | 1.5460 | 7.266 | 0.0001 | 1.0e-07 | Unconditional enrichment |
| Endothelial mural | 0.7551 | -3.408 | 1.000 | 1.000 | Unconditional enrichment |
| Interneurons | 0.7096 | -4.418 | 1.000 | 1.000 | Unconditional enrichment |
| Microglia | 0.1491 | 5.769 | 0.0001 | 1.0e-07 | Unconditional enrichment |
| Oligodendrocytes | 1.0878 | 1.170 | 0.1268 | 0.2657 | Unconditional enrichment |
| Pyramidal CA1 neurons | 0.7164 | -5.011 | 1.000 | 1.000 | Unconditional enrichment |
| Pyramidal SS neurons | 0.6804 | -5.831 | 1.000 | 1.000 | Unconditional enrichment |
| Proteomics | **dataset** |  |  |  |  |
| Astrocytes (ependymal) | 1.1626 | 2.0493 | 0.0272 | 0.0952 | Unconditional enrichment |
| Endothelial mural | 0.6887 | -4.103 | 1.000 | 1.000 | Unconditional enrichment |
| Interneurons | 0.9915 | -0.1187 | 0.5307 | 0.7921 | Unconditional enrichment |
| Microglia | 1.0772 | 0.8521 | 0.1941 | 0.4530 | Unconditional enrichment |
| Oligodendrocytes | 1.1988 | 2.4744 | 0.0100 | 0.0700 | Unconditional enrichment |
| Pyramidal CA1 neurons | 0.9701 | -0.4914 | 0.6789 | 0.7921 | Unconditional enrichment |
| Pyramidal SS neurons | 0.9732 | -0.4548 | 0.6703 | 0.7921 | Unconditional enrichment |
| GWAS-orthologs | **dataset** |  |  |  |  |
| Astrocytes (ependymal) | 1.2336 | 1.7514 | 0.0480 | 0.3360 | Unconditional enrichment |
| Endothelial mural | 0.8842 | -0.8969 | 0.8124 | 0.9478 | Unconditional enrichment |
| Interneurons | 0.9580 | -0.3519 | 0.6150 | 0.8610 | Unconditional enrichment |
| Microglia | 1.0944 | 0.6165 | 0.2583 | 0.8165 | Unconditional enrichment |
| Oligodendrocytes | 1.0343 | 0.2531 | 0.3668 | 0.8165 | Unconditional enrichment |
| Pyramidal CA1 neurons | 1.0041 | 0.0397 | 0.4666 | 0.8165 | Unconditional enrichment |
| Pyramidal SS neurons | 0.7883 | -2.1317 | 0.9908 | 0.9908 | Unconditional enrichment |
| Combined AD multi-omics | **dataset** |  |  |  |  |
| Astrocytes (ependymal) | 1.3568 | 6.5462 | 0.0001 | 1.0e-07 | Unconditional enrichment |
| Endothelial mural | 0.7616 | -4.567 | 1.000 | 1.000 | Unconditional enrichment |
| Interneurons | 0.8652 | -2.821 | 0.9986 | 1.000 | Unconditional enrichment |
| Microglia | 1.1979 | 3.1434 | 0.0012 | 0.0056 | Unconditional enrichment |
| Oligodendrocytes | 1.1137 | 2.0620 | 0.0237 | 0.0737 | Unconditional enrichment |
| Pyramidal CA1 neurons | 0.8876 | -2.7150 | 0.9976 | 1.000 | Unconditional enrichment |
| Pyramidal SS neurons | 0.8344 | -4.1285 | 1.000 | 1.000 | Unconditional enrichment |
| Astrocytes (ependymal) | 1.3797 | 7.5402 | 0.0001 | 1.0e-07 | Conditional enrichment (microglia controlled) |
| Endothelial mural | 0.7668 | -4.7705 | 1.000 | 1.000 | Conditional enrichment (microglia controlled) |
| Interneurons | 0.9222 | -1.8281 | 0.9695 | 1.000 | Conditional enrichment (microglia controlled) |
| Microglia | 0.9998 | -0.3555 | 0.6367 | 1.000 | Conditional enrichment (microglia controlled) |
| Oligodendrocytes | 1.1315 | 2.5311 | 0.0099 | 0.03885 | Conditional enrichment (microglia controlled) |
| Pyramidal CA1 neurons | 0.9359 | -1.6967 | 0.9562 | 1.000 | Conditional enrichment (microglia controlled) |
| Pyramidal SS neurons | 0.8802 | -3.3302 | 0.9999 | 1.000 | Conditional enrichment (microglia controlled) |
| Astrocytes (ependymal) | 0.9995 | -1.5705 | 0.9424 | 1.000 | Conditional enrichment (astrocyte controlled) |
| Endothelial mural | 0.8188 | -3.7026 | 0.9999 | 1.000 | Conditional enrichment (astrocyte controlled) |
| Interneurons | 0.9439 | -1.2257 | 0.8891 | 1.000 | Conditional enrichment (astrocyte controlled) |
| Microglia | 1.2559 | 4.4759 | 0.0001 | 1.0e-07 | Conditional enrichment (astrocyte controlled) |
| Oligodendrocytes | 1.1245 | 2.3867 | 0.0111 | 0.03885 | Conditional enrichment (astrocyte controlled) |
| Pyramidal CA1 neurons | 0.9625 | -0.9519 | 0.8268 | 1.000 | Conditional enrichment (astrocyte controlled) |
| Pyramidal SS neurons | 0.9185 | -2.1752 | 0.9863 | 1.000 | Conditional enrichment (astrocyte controlled) |
| Astrocytes (ependymal) | 1.3459 | 6.7564 | 0.0001 | 1.0e-07 | Conditional enrichment (oligodend. controlled) |
| Endothelial mural | 0.7802 | -4.4094 | 1.000 | 1.000 | Conditional enrichment (oligodend. controlled) |
| Interneurons | 0.9034 | -2.2123 | 0.9884 | 1.000 | Conditional enrichment (oligodend. controlled) |
| Microglia | 1.1907 | 3.2431 | 0.0008 | 0.00448 | Conditional enrichment (oligodend. controlled) |
| Oligodendrocytes | 1.0000 | 0.0919 | 0.4409 | 1.000 | Conditional enrichment (oligodend. controlled) |
| Pyramidal CA1 neurons | 0.8928 | -2.6396 | 0.9969 | 1.000 | Conditional enrichment (oligodend. controlled) |
| Pyramidal SS neurons | 0.8594 | -3.5656 | 1.000 | 1.000 | Conditional enrichment (oligodend. controlled) |

SD refers to Standard Deviation, B-H adj. refers to Benjamini-Hochberg (B-H) FDR-adjusted p-value.
